# Observed-to-Expected Fetal Losses Following mRNA COVID-19 Vaccination in Early Pregnancy

**DOI:** 10.1101/2025.06.18.25329352

**Authors:** Josh Guetzkow, Tal Patalon, Sivan Gazit, Tracy Beth Høeg, Joseph Fraiman, Yaakov Segal, Retsef Levi

**Affiliations:** Department of Sociology & Anthropology, Institute of Criminology Hebrew University of Jerusalem Jerusalem, Israel 91905; Maccabi Research and Innovation Center, Maccabi Healthcare Services Tel Aviv-Yafo, Israel 68012, Arison School of Business, Reichman University Herzliya, Israel 46101; Maccabi Research and Innovation Center, Maccabi Healthcare Services Tel Aviv, Israel 68012; Sloan School of Management, Massachusetts Institute of Technology Department of Emergency Medicine, University of California - San Francisco Department of Clinical Research, University of Southern Denmark Cambridge, MA USA 02142; Baromedical Research Institute Harvey, LA USA 70058; Health Division, Maccabi Healthcare Services Tel Aviv, Israel 68012; Sloan School of Management, Massachusetts Institute of Technology Cambridge, MA USA 02142

## Abstract

**Background:** The clinical trials used to approve COVID-19 vaccines excluded pregnant women, and existing safety assessments of COVID-19 vaccination, particularly during early stages of pregnancy, are limited to observational studies prone to various types of potential bias, including healthy vaccinee bias.

**Methods:** The study includes pregnancies in Israel with last menstruation period (LMP) between March 1, 2016 and February 28, 2022. The main analysis presents observed-to-expected comparisons of the number of eventual fetal losses among pregnant women exposed to mRNA COVID-19 vaccination (almost all Pfizer) during gestational weeks 8-13 and 14-27, respectively. Women vaccinated for influenza during gestational weeks 8-27, as well as women vaccinated prior to pregnancy for COVID-19 or influenza, were used as comparative controls. Cohort-specific expected number of fetal losses are established based on estimates from a regression model trained on historical data from 2016-2018 that incorporates individual-level risk factors and gestational week of each pregnant woman included in the cohort.

**Results:** Analysis of 226,395 singleton pregnancies in Israel from 2016 to 2022 indicates that COVID-19 vaccination with dose 1 during weeks 8-13 was associated with *higher-than-expected* observed number of fetal losses of approximately 13 versus 9 expected for every 100 exposed pregnancies, i.e., nearly 3.9 (95% CI: [2.55-5.14]) additional fetal losses above expected per 100 pregnancies Most of the excess fetal losses occurred after gestational week 20 and nearly half occurred after gestational week 25. Similarly, women vaccinated with dose 3 during weeks 8-13 exhibited a *higher-than-expected* number of fetal losses with nearly 1.9 (95% CI: 0.39-3.42]) additional fetal losses above expected per 100 pregnancies. In contrast, pregnant women vaccinated for influenza during weeks 8-27 exhibited a consistently *lower-than-expected* observed number of fetal losses, likely the result of healthy vaccinee bias. Women vaccinated for COVID-19 or influenza prior to pregnancy exhibited *according-to-expected* or *lower-than-expected* numbers of fetal losses.

**Conclusion:** The results provide evidence for a substantially *higher-than-expected* number of eventual fetal losses associated with COVID-19 vaccination during gestational weeks 8-13.

## INTRODUCTION

Pregnant women were excluded from the pivotal randomized clinical trials used for the initial regulatory approvals of the COVID-19 vaccines. The one subsequent randomized clinical trial in pregnant women was conducted by Pfizer and only included 173 women who were vaccinated in relatively advanced stages of pregnancy (gestational weeks 24-34) (Pfizer 2023).

Observational studies on COVID-19 vaccination during pregnancy predominantly compare outcomes between vaccinated and unvaccinated pregnant women during active vaccination campaigns using various regression models (Fell, Dhinsa, et al. 2022, Magnus, et al. 2022, Calvert, Carruthers, et al. 2022, Velez, et al. 2023, Calvert, Carruthers, et al. 2023, Goldshtein, Steinberg, et al. 2022, Hui, et al. 2023, Rimmer, et al. 2023). Among these studies, only a few have considered the impact of COVID-19 vaccination in early stages of pregnancy, when the risk of teratogenicity is likely the highest (Calvert, Carruthers, et al. 2022, Velez, et al. 2023, Shimabukuro, et al. 2021). Additional studies of the association between COVID-19 vaccination in early pregnancy and spontaneous abortions (prior to week 20) used a case control method. (Kharbanda, Haapala and DeSilva, et al. 2021, Kharbanda, Haapala and Lipkind, et al. 2023)

With a few exceptions (Aharon, et al. 2022, Kuhbandner and Reitzner 2023, Velez, et al. 2023), studies on COVID-19 vaccination during pregnancy have not reported on any statistically significant association of adverse pregnancy outcomes with COVID-19 vaccination during pregnancy. (Rimmer, et al. 2023, Prasad, et al. 2022)

However, these observational studies have known methodological limitations related to controlling for gestational timing of vaccination, the inclusion and exclusion criteria applied, and potential confounding biases that are not controlled for. (Fell, Dimitris, et al. 2021) In particular, observed and unobserved differences in health status between vaccinated and unvaccinated women could affect the studied outcomes and be a source of methodological bias, referred to as *healthy (or unhealthy) vaccinee bias*. (Høeg, Duriseti and Prasad 2023, Remschmidt, Wichmann and Harder 2015)

The present study aimed to assess the impact of COVID-19 vaccination during gestational weeks 8-27 by leveraging anonymized data from Maccabi Healthcare Services (MHS), the second largest health fund in Israel. The main analysis considered different exposed cohorts of pregnant women, defined based on the timing and dose of vaccination, and using historical data, analyzed the respective observed-to-expected difference in the number of fetal losses within each cohort.

Israel was the first country in the world to launch a nationwide COVID-19 vaccination campaign, starting in December 2020, using almost exclusively the Pfizer BNT162b2 mRNA vaccine. In August 2021, Israel was again the first country to vaccinate the population with a booster dose. A recommendation to vaccinate pregnant women was published by the Israeli Ministry of Health on January 19, 2021, and was initially restricted to the 2^nd^ and 3^rd^ trimesters before being extended to all stages of pregnancy on February 1, 2021. There are several studies on the association between COVID-19 vaccination and various pregnancy outcomes based on data from Israel, but they share similar methodological limitations to those mentioned above. (Wainstock, Yoles and Sergienko 2021, Lipschuetz, et al. 2023, Dick, et al. 2022, Rottenstreich, et al. 2022, Goldshtein, Steinberg, et al. 2022)

## MATERIALS AND METHODS

### Data Sources

This study relies on the centralized electronic health record (EHR) database maintained by MHS since the 1990’s, comprised of a longitudinal database on 2.7 million patients (26% of Israel’s population) with minimal turnover (∼1% annually). Anonymized data from the MHS pregnancy registry were extracted on May 6, 2023, at which point the registry contained information on 1,330,845 pregnancies since 1990. This study used 226,395 pregnancies with complete and reliable data and LMP between March 1, 2016 and February 28, 2022. The registry includes information routinely recorded by obstetricians, including pregnancy outcomes and the date on which they occurred, date of last menstrual period (LMP), number of embryos, and whether the pregnancy was considered high risk, along with the date on which the pregnancy entered the high-risk category. Data from the pregnancy registry were joined with other EHR data pertaining to diagnosed clinical conditions, claims for medical services and procedures, and a specialized COVID-19 registry that included information on COVID-19 vaccination records and laboratory findings from a central laboratory. MHS’ institutional review board approved the study, which was exempt from informed consent due to its retrospective nature and the use of routinely collected anonymized information.

### Exposures

The primary analysis included cohorts of pregnant women exposed to dose 1 or 3 of any COVID-19 vaccination during pregnancy, with separate cohorts for exposure during gestational weeks 8-13 and weeks 14-27, respectively. Dose 2 was included only as a supplementary analysis, since the vast majority of women who received dose 1 continued to receive dose 2. The dose 3 cohorts include only women who received two doses prior to pregnancy. The study also included single-week cohorts for gestational weeks W=8,…,27, each composed of women who vaccinated on the specific gestational week W for COVID-19 (dose 1 or 3), respectively. The prospective risk of each woman included in these COVID-19 vaccination cohorts is assessed prospectively from a *risk week* defined as the gestational week of vaccination. (See the discussion regarding the outcomes below.)

Women vaccinated during gestational weeks 1-7 were excluded, because identification and follow-up of pregnancies during these weeks is partial and inconsistent, raising concerns about reporting biases; for example, vaccinated women might be more prone to report early fetal loss. By week 8 or 9, almost all pregnancies are already under follow-up and documentation is substantially more consistent.

To address, at least partially, bias from potentially unobserved confounding covariates, the analysis considered two types of comparative control cohorts. First, control cohorts were similarly defined for exposure to influenza vaccination during gestational weeks 8-13 and 14-27, as well as single-weeks W=8,…,27. Second, exposure to vaccination prior to pregnancy was considered as another control with single-week cohorts for gestational weeks W=8,…,27, each including women still pregnant at the start of week W, who received doses 1 and 2, dose 3 or influenza vaccination prior to pregnancy. In these latter cohorts, the risk week is defined as the respective gestational week W.

Exploratory analysis included exposure to SARS-CoV-2 infection during pregnancy (confirmed by positive PCR test), with cohorts corresponding to infection occurring during gestational weeks 8-13 and 17-24 with and without prior COVID-19 vaccination, respectively.

### Outcomes

The main outcome of the study was eventual fetal loss throughout pregnancy, which is a composite measure that includes spontaneous and induced abortions and stillbirths. The composite outcome was determined based on the pregnancy registry, diagnosis and procedural codes. (See Table S1 in the Supplementary Appendix for detailed description.) While spontaneous abortions and stillbirths occur involuntarily and are driven by biological mechanisms, induced abortions can be either elective or therapeutic (i.e., driven by medical reasons). Elective abortions are typically driven by personal considerations or reasons such as the age of the mother and the circumstances of the pregnancy (e.g., outside marriage or the result of relationships prohibited by criminal law). Medical reasons for induced abortions (e.g., fetal defects and malformation) involve biological mechanisms, overlapping with those of spontaneous abortions and stillbirths. Diagnosis and claims data related to induced abortions often do not include clear documentation of the underlying rationale and many are driven by fetal health concerns.

To assess the plausibility that the findings regarding the eventual fetal loss rates were not entirely driven by elective decisions of women, late fetal losses from gestational weeks 14, 20 and 25, respectively, were considered as additional outcomes. In Israel, all induced abortions must be pre-approved by special committees that are appointed within healthcare institutions. A recent report by the Israeli Ministry of Health on temporal trends of induced abortions during 1990-2022 indicates that elective abortions after gestational week 14 have become increasingly rare, and induced abortions after week 24 are rare and only granted for medical reasons. (Israel Ministry of Health 2023)

### Observed-to-Expected Numbers of Eventual Fetal Loss

The primary analyses in this study followed a commonly used approach in vaccine safety studies where the observed incidence of adverse events among vaccinated individuals is compared to the expected incidence based on background rates in the general population. (Gordillo-Marañón, et al. 2024, Mahaux, Bauchau and Holle 2015) Unlike existing studies on the safety of vaccination during pregnancy that have used expected background incidence rates that are not specific to the study’s population (Shimabukuro, et al. 2021), the current study estimated the cohort-specific expected number of fetal losses adjusted for the individual-level characteristics of the vaccinated women.

The adjusted estimates of the expected number of fetal losses were calculated based on an individual-level *baseline reference model* obtained by training a pooled logistic regression model on pregnancies with LMPs between March 1, 2016 and February 28, 2018. This model took as an input a pregnancy on a given gestational risk week. It then estimated the prospective risk of eventual fetal loss from that week throughout the pregnancy, adjusting for the gestational age and calendar month of the pregnancy, age of the woman, as well as multiple covariates that capture the woman’s health status, health-seeking behavior and other socioeconomic factors. For a detailed description of the baseline reference model, covariates and model results see Supplementary Appendix Section S1 and Tables S1 and S2.

The cohort-specific expected number of eventual fetal losses was calculated by summing the predicted individual risk probabilities of all the women in the cohort, which were calculated based on the week in which each of them were exposed (i.e., the risk week). The expected number was then compared to the observed number of fetal losses in the cohort. Figure S1 in the Supplementary Appendix illustrates how the analysis was conducted on the vaccination cohorts for gestational weeks 8-13. The cohort-specific expected and observed numbers of fetal losses are reported (as number of fetal losses per 100 pregnancies), together with the observed-to-expected difference, the 95% confidence interval (CI), and the number of pregnancies included in each cohort. The ratios of observed-to-expected fetal losses with corresponding 95% CI’s are reported separately in the Supplementary Appendix. For a description of how the expected values and CI’s were calculated, see section S2 in the Supplementary Appendix.

The analysis can be interpreted as a simulated trial, where each vaccinated pregnant woman included in the cohort was ‘matched’ with a ‘*synthetic* unvaccinated control’ with similar individual and pregnancy characteristics. The outcomes of the synthetic controls were ‘simulated’ based on the baseline reference model and compared to the observed outcomes. By comparing vaccinated women to historical (synthetic) controls rather than controls within the vaccination period, the observed-to-expected analysis allowed maximal ‘matching’ on observed covariates and uncensored follow-up time.

#### Late fetal losses

To address the potential concern that women vaccinated for COVID-19 (dose 1 or 3) or influenza in early gestational weeks (8-13) could have been more (or less) prone to have purely elective induced abortions, the observed-to-expected difference in the number of eventual fetal losses was calculated from weeks 14, 20 and 25 for women vaccinated in weeks 8-13. Notably, the individual risk of eventual fetal loss, for each woman in these cohorts, was estimated prospectively at gestational weeks 14, 20 and 25, separately, for all the women who were still pregnant at the beginning of the respective week. In addition, the percentage of women experiencing fetal loss from gestational week 25 onward was calculated for women receiving COVID-19 vaccination (dose 1 or 3) or influenza vaccination, in each of the weeks W=8,…,13. These were then compared to the same percentage among all women still pregnant at the start of each week W.

### Validation and Robustness Analyses

To obtain out-of-sample validation of the baseline reference estimates and assess how consistent they were between different analysis periods, the observed-to-expected analysis was conducted for influenza vaccination cohorts with LMPs from March 1, 2018 to February 28, 2019. These included the cohorts of gestational weeks 8-13, weeks 14-27, as well as single-week cohorts W=8,…,27 with influenza vaccination prior to pregnancy but in the same influenza season as the LMP.

Several additional robustness analyses were conducted. First, the main observed-to-expected analyses were repeated using a baseline reference model that was estimated on data from March 1, 2016 through February 28, 2019. Second, to assess the sensitivity of the results to the choice of the follow-up start time (week 8), the observed-to-expected analysis was repeated when pregnancies were followed from week 10.

Complementary analyses were conducted to compare both the risk score and covariate distributions of the cohort of women vaccinated for COVID-19 in weeks 18-13 with the corresponding influenza vaccination cohort, as well as women exposed to influenza or COVID-19 vaccination prior to pregnancy. To assess the correlation between influenza and COVID-19 vaccination in early stages of pregnancy, the women whose pregnancy’s timing allowed them to receive dose 1 of the COVID-19 vaccine during gestational weeks 8-13 and 8-27, respectively, were identified. Within these groups, women who received influenza vaccines in the same season were compared to women who did not.

## RESULTS

Figure 1 presents the pregnancies included and excluded from the analysis, along with the reasons for exclusion. Table S3 presents descriptive statistics on the covariates for women included in each of the three analysis periods (Reference, Validation and COVID-19). It shows small differences in the mean value of some covariates, which were adjusted for in the observed-to-expected analyses. The rates of COVID-19 vaccination during pregnancy and their calendar timing are described in the Supplementary Appendix section S4 and Figure S5.

**Figure 1.**
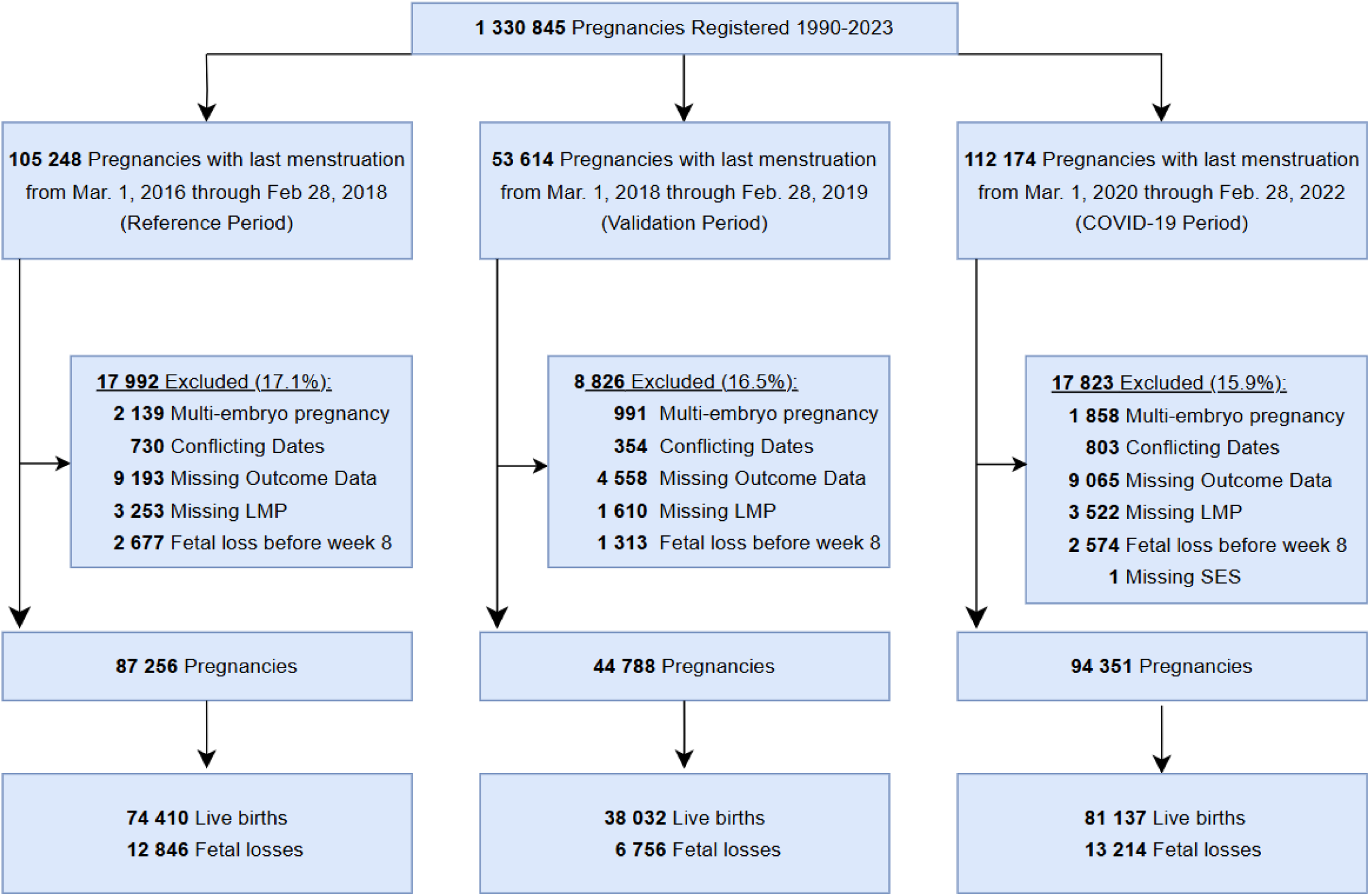
Pregnancies Included and Excluded from Analyses.

### Observed-to-Expected Analysis

#### Vaccination during pregnancy

Table 1 shows the results of the observed-to-expected analysis for women vaccinated during weeks 8-13 and 14-27. Women vaccinated for COVID-19 with doses 1 or 3 during weeks 8-13 exhibited *higher-than-expected* numbers of observed fetal losses throughout pregnancy. The observed-to-expected differences were 3.85 (95% CI: [2.55-5.14]) for dose 1, and 1.9 (95% CI: 0.39-3.42]) for dose 3, i.e., an additional 3.85 and 1.9 observed fetal losses above expected per 100 pregnancies, respectively. For COVID-19 vaccination during weeks 14-27, the observed numbers of fetal losses were *lower-than-expected*, with observed-to-expected differences of −0.79 (95% CI: [−1.07 --0.51]) for dose 1 and −0.85 (95% CI: [−0.48 --0.37]) for dose 3.

**Table 1.**
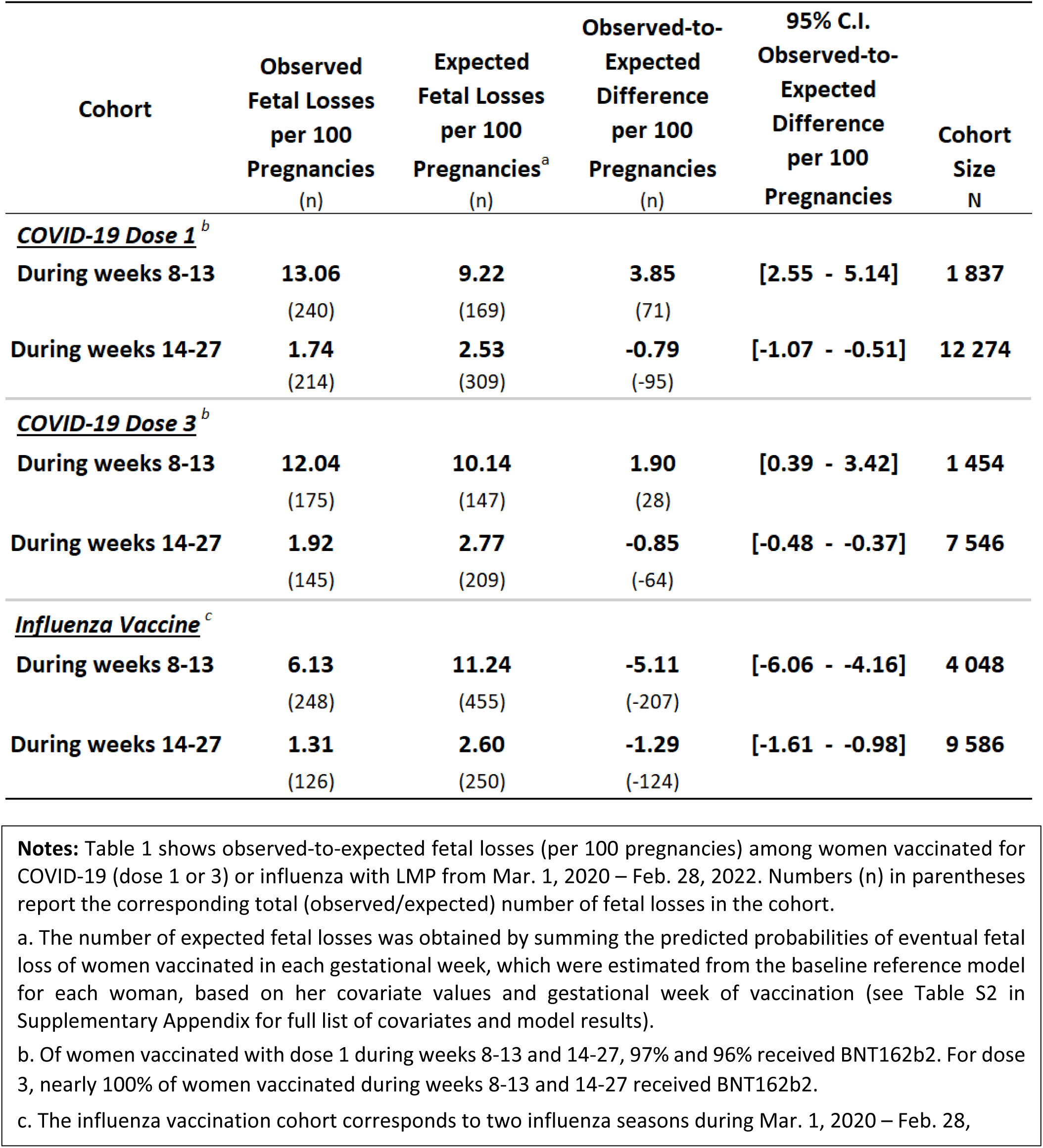
Observed-to-Expected Eventual Fetal Losses among Women Vaccinated for COVID-19 or Influenza in Gestational Weeks 8-13 and 14-27.

In marked contrast, the corresponding influenza vaccination cohorts exhibited *lower-than-expected* numbers of observed fetal losses with observed-to-expected differences of −5.11 (95% CI: [−6.06 --4.16] and −1.24 (95% CI: [−1.61 --0.98]) for weeks 8-13 and 14-27, i.e., 5.11 and 1.24 fewer fetal losses per 100 pregnancies compared to expected, respectively. The results for COVID-19 vaccine dose 2 are similar to dose 1 and are shown in Table S4 in the Supplementary Appendix. The observed-to-expected fetal loss ratios are reported in Table S6.

Figure 2 shows the observed-to-expected results for each individual week-level cohort W=8,…,27, for COVID-19 dose 1 and influenza vaccination. These exhibited patterns consistent with the results in Table 1 for cohorts of women who vaccinated during weeks 8-13 and 14-27. Figure 2(a) shows that, for each of the weeks W=8,…,13, women who were vaccinated with dose 1 during week W, exhibited *higher-than-expected* numbers of observed fetal losses with observed-to-expected differences as high as an additional 9 fetal losses over expected per 100 pregnancies. The observed-to-expected difference remained positive but decreased from week 9 through week 13, and in weeks W=14,…,27, the pattern was reversed with a *lower-than-expected* number of observed fetal losses. In contrast, the corresponding influenza vaccination cohorts shown in Fig. 2(b) exhibited substantially *lower-than-expected* observed numbers of fetal loss rates throughout all weeks W=8,…,27. The results for COVID-19 vaccination doses 2 and 3 are presented in Figure S2 in the Supplementary Appendix.

**Figure 2.**
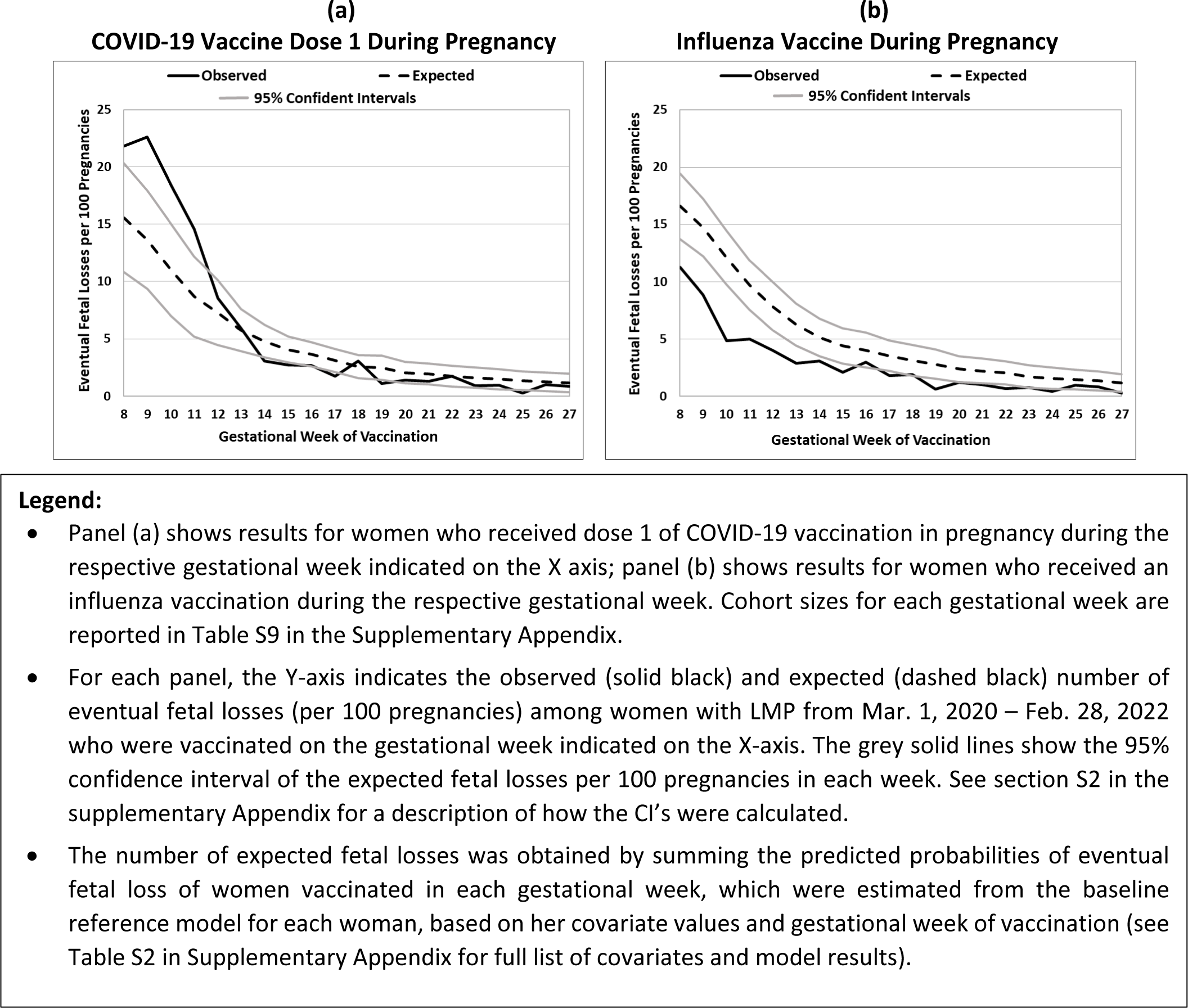
Observed-to-Expected Eventual Fetal Losses by Gestational Week of Vaccination among Women Vaccinated for COVID-19 or Influenza.

#### Late fetal losses

Table 2 shows the results of the observed-to-expected eventual fetal loss analysis of the women who vaccinated for COVID-19 (dose 1 or 3) or influenza during gestational weeks 8-13 and were still pregnant at the beginning of gestational weeks 14, 20 and 25, respectively. Both the dose 1 and 3 cohorts continued to exhibit substantial residual *higher-than-expected* numbers of eventual fetal losses through gestational week 25. For dose 1, the observed-to-expected difference in week 14 was 3.05 (95% CI: [2.06 – 4.05]), i.e., more than 3 additional fetal losses per 100 pregnancies over expected, which was very close to the 3.85 observed-to-expected difference reported in Table 1. The observed-to-expected differences in weeks 20 and 25 were 2.47 (95% CI: [1.78 – 3.16]) and 1.66 (95% CI: [1.11 – 2.]) fetal losses above expected per 100 pregnancies, respectively. For dose 3, the observed-to-expected difference in week 14 was 2.73 (95% CI: [1.57 – 3.89]) fetal losses above expected per 100 pregnancies, which was higher than the 1.9 reported in Table 1, and in weeks 20 and 25, it was 1.52 (95% CI: [0.71 – 2.54]) and 0.95 (95% CI: [0.30 – 1.]) fetal losses above expected per 100 pregnancies, respectively. In contrast, the *lower-than-expected* observed number of eventual fetal losses of the influenza cohort exhibited rapidly shrinking negative observed-to-expected differences with −2.27 (95% CI: [−2.96 --1.58]), −0.91 (95% CI: [−1.39 --0.44]) and −0.78 (95% CI: [−1.16 --0.39]) fetal losses per 100 pregnancies in weeks 14, 20 and 25, respectively, compared to the −5.11 fetal losses per 100 pregnancies reported in Table 1. (See Table S6 for the corresponding observed-to-expected fetal loss ratios.) Furthermore, Table 3 shows that 2.72%, 1.79% and 0.72% of the women who vaccinated during gestational weeks 8-13, with dose 1, dose 3 and influenza, respectively, had a fetal loss from gestational week 25 onward, compared to 0.99% and 1.09% for all women who were still pregnant in weeks 8 and 13, respectively.

**Table 2.** Observed-to-Expected Eventual Late Fetal Losses from Weeks 14, 20 and 25 among Women Vaccinated for COVID-19 or Influenza in Gestational Weeks 8-13.

| Cohort | Observed<br>Fetal Losses<br>per 100<br>Pregnancies<br>(n) | Expected<br>Fetal Losses<br>per 100<br>Pregnancies<sup>a</sup><br>(n) | Observed-to-Expected<br>Difference<br>per 100<br>Pregnancies<br>(n) | 95% C.I.<br>Observed-to-Expected<br>Difference<br>per 100<br>Pregnancies | Cohort<br>Size<br>N |
| --- | --- | --- | --- | --- | --- |
| **<u>COVID-19 Dose 1<sup>b</sup></u>** |  |  |  |  |  |
| *From Week 14* | **7.85**<br>(136) | **4.79**<br>(83) | **3.05**<br>(53) | **[2.06 - 4.05]** | **1 733** |
| *From Week 20* | **4.60**<br>(77) | **2.13**<br>(36) | **2.47**<br>(41) | **[1.78 - 3.16]** | **1 674** |
| *From Week 25* | **3.04**<br>(50) | **1.37**<br>(23) | **1.66**<br>(27) | **[1.11 - 2.23]** | **1 646** |
| **<u>COVID-19 Dose 3<sup>b</sup></u>** |  |  |  |  |  |
| *From Week 14* | **7.92**<br>(110) | **5.19**<br>(72) | **2.73**<br>(38) | **[1.57 - 3.89]** | **1 389** |
| *From Week 20* | **3.83**<br>(51) | **2.31**<br>(31) | **1.52**<br>(20) | **[0.71 - 2.54]** | **1 330** |
| *From Week 25* | **2.44**<br>(32) | **1.49**<br>(19) | **0.95**<br>(13) | **[0.30 - 1.61]** | **1 311** |
| **<u>Influenza Vaccine<sup>c</sup></u>** |  |  |  |  |  |
| *From Week 14* | **3.01**<br>(118) | **5.28**<br>(207) | **-2.27**<br>(-89) | **[-2.96 - -1.58]** | **3 918** |
| *From Week 20* | **1.45**<br>(56) | **2.36**<br>(91) | **-0.91**<br>(-35) | **[-1.39 - -0.44]** | **3 856** |
| *From Week 25* | **0.76**<br>(29) | **1.53**<br>(59) | **-0.78**<br>(-30) | **[-1.16 - -0.39]** | **3 828** |

**Notes:** Table 2 shows the number of observed-to-expected eventual fetal losses (per 100 pregnancies) for women with LMP from Mar. 1, 2020 – Feb. 28, 2022 who were vaccinated for COVID-19 (dose 1 or 3) or influenza during gestational weeks 8-13. Numbers (n) in parentheses report the corresponding total number of (observed/expected) fetal losses in the cohort.
a. The number of expected fetal losses was obtained by summing the predicted probability of eventual fetal loss of women in each vaccination cohort from the respective gestational week (14, 20, 25), which were estimated for each woman from the reference baseline model, based on her covariate values and gestational week of pregnancy (14, 20, 25) (see Table S2 in Supplementary Appendix for list of covariates and model results). Women who experienced fetal loss prior to the respective week (14, 20, 25) were not included in the calculation.
b. Of women vaccinated with dose 1 during weeks 8-13, 97% received BNT162b2. For dose 3, nearly 100% of women vaccinated during weeks 8-13 received BNT162b2.
c. The influenza vaccination cohort corresponds to two seasons from Mar. 1, 2020 – Feb. 28, 2022.

**Table 3.** Observed Fetal Losses from Week 25 among Women Vaccinated for COVID-19 or Influenza and All Women in Gestational Weeks 8-13.

| **Week** | **Cohort Size**<br>**(N)** | **Fetal Losses**<br>**from Week 25**<br>**(n)** | **Fetal Losses**<br>**from Week 25**<br>**(%)** |
| --- | --- | --- | --- |
| **DOSE 1** |  |  |  |
| 8 | 211 | 5 | 2.37% |
| 9 | 239 | 9 | 3.77% |
| 10 | 227 | 5 | 2.20% |
| 11 | 240 | 8 | 3.33% |
| 12 | 315 | 8 | 2.54% |
| 13 | 605 | 15 | 2.48% |
| 8-13 | 1837 | 50 | 2.72% |
| **DOSE 3** |  |  |  |
| 8 | 205 | 2 | 0.98% |
| 9 | 231 | 4 | 1.73% |
| 10 | 218 | 7 | 3.21% |
| 11 | 230 | 8 | 3.48% |
| 12 | 309 | 3 | 0.97% |
| 13 | 590 | 8 | 1.36% |
| 8-13 | 1783 | 32 | 1.79% |
| **INFLUENZA** |  |  |  |
| 8 | 621 | 3 | 0.48% |
| 9 | 736 | 6 | 0.82% |
| 10 | 706 | 2 | 0.28% |
| 11 | 700 | 10 | 1.43% |
| 12 | 621 | 4 | 0.64% |
| 13 | 664 | 4 | 0.60% |
| 8-13 | 4048 | 29 | 0.72% |
| **ALL WOMEN** |  |  |  |
| 8 | 94351 | 935 | 0.99% |
| 9 | 92492 | 935 | 1.01% |
| 10 | 90096 | 935 | 1.04% |
| 11 | 88093 | 935 | 1.06% |
| 12 | 86677 | 935 | 1.08% |
| 13 | 85567 | 935 | 1.09% |

**Notes:** Table 3 panels show the rate of eventual fetal loss beginning from week 25 for women vaccinated for COVID-19 dose 1, dose 3, influenza and for all women with LMP from Mar. 1, 2020 – Feb. 28, 2022. For vaccination cohorts, the N is the number of women vaccinated in the given week, and for all women the N is the number of women still pregnant at the start of the given week. The vaccination cohorts also show combined calculations for all women in the 8-13 week cohorts, which can be compared to week 13 for all women.

#### Vaccination prior to pregnancy

Figure 3(a) shows observed-to-expected results for each week W=8,…,27 among women who received two doses prior to pregnancy, whose pregnancies survived to the start of the respective week W and who had not vaccinated with a third dose by the start of the week. Figure 3(b) shows the same for women vaccinated for influenza in the same season as their LMP but prior to pregnancy. Unlike results for vaccination during pregnancy, both COVID-19 and influenza cohorts had similar observed and expected numbers of fetal losses, and the observed numbers were *according-to-expected* or slightly *lower-than-expected*. Figure S3 in the Supplementary Appendix shows similar results for women vaccinated with 3 doses prior to pregnancy.

#### SARS-CoV-2 infections

The observed numbers of fetal losses were slightly *lower-than-expected* for women with SARS-CoV-2 infections during gestational weeks 8-13 and 14-27 before and after COVID-19 vaccination, except for SARS-CoV-2 infections in gestational weeks 8-13 among unvaccinated women, but all the 95% CIs of the respective observed-to-expected differences included 0 (Table S8). SARS-CoV-2 infection rates before and during pregnancy are described in the Supplementary Appendix section S4.

### Validation and Robustness Results

The results of the observed-to-expected analysis for influenza vaccination during the pre-pandemic period from March 1, 2018 – February 28, 2019 were very similar to the results of the primary analysis, suggesting that year-to-year changes in patterns of the observed-to-expected differences in the numbers of eventual fetal losses were relatively low (Table S5, Figure S4).

Comparison between women in the dose 1 during gestational weeks 8-13 cohort to the control cohorts (influenza vaccination during weeks 8-13 and COVID-19 and influenza vaccination prior to pregnancy) indicates that the risk scores of women in the control cohorts were relatively higher (Table S7). This could be explained by differences in covariate distributions, where the control cohorts had a higher fraction of women with comorbidities and risk factors.

Most women (89%) who vaccinated with dose 1 during gestational weeks 8-27 had an LMP from Oct. 2020 – Jan. 2021 (see Figure S5 for monthly distribution). Overall, 20,383 pregnancies with LMPs during these months had an opportunity to receive dose 1 of COVID-19 vaccination in gestational weeks 8-27. Of those, 2,984 had received an influenza vaccine during the 2020-2021 influenza season. Women who received influenza vaccine were about 2-fold more likely to receive dose 1 of the COVID-19 vaccine than women who did not (15.08% vs. 7.25% for weeks 8-13 and 71.31% vs. 37.94% for weeks 8-27).

Results with the baseline reference model estimated on data from March 2016 through February 2019 were very similar to the results of the primary analysis. The results also remained very similar when pregnancies were followed from week 10.

## DISCUSSION

The main finding of this study is that COVID-19 vaccination with any dose during gestational weeks 8-13 of pregnancy was associated with *higher-than-expected* observed number of fetal losses (Tables 1, S4). For dose 1, this amounted to close to 4 (3.85) additional fetal losses above expected for every 100 pregnancies that were exposed to dose 1 of COVID-19 vaccination during weeks 8-13, or approximately 13 observed fetal losses per 100 pregnancies versus 9 expected. Similarly, women vaccinated with dose 3 during weeks 8-13 exhibited nearly 1.9 additional fetal losses per 100 pregnancies over expected.

Women who were vaccinated during weeks 8-13 sustained *higher-than-expected* observed numbers of eventual fetal losses over the course of their pregnancy. Out of the 3.85 additional fetal losses per 100 pregnancies above expected for dose 1 (Table 1), over 3 occurred from week 14 onward, and slightly under half, or 1.66 fetal losses, occurred from gestational week 25 onward (Table 2). Moreover, 2.72% of women vaccinated with dose 1 and 1.79% of women vaccinated with dose 3 during weeks 8-13 had a fetal loss from week 25 onward, which was substantially higher than the rate of 1% for all pregnant women and 0.79% of women vaccinated for influenza during (Table 3). This indicates that a large part of the *higher-than-expected* observed number of fetal losses associated with COVID-19 vaccination during gestational weeks 8-13 unambiguously stemmed from late fetal losses, i.e., stillbirths and spontaneous or therapeutic abortions driven by biological mechanisms and medical reasons, rather than to behavioral patterns that typically drive purely elective abortions.

Comparing the observed-to-expected patterns of COVID-19 vaccination during weeks 8-13 to the corresponding patterns during weeks 14-27 as well as to the comparative control cohorts provides important context to their interpretation. The *higher-than-expected* observed number of fetal losses for COVID-19 vaccination during weeks 8-13 changed rapidly, and gestational weeks 14-27 exhibited *lower-than-expected* observed numbers. Additionally, in contrast to COVID-19 vaccination, influenza vaccination during pregnancy exhibited substantial *lower-than-expected* numbers of observed fetal losses throughout gestational weeks 8-27, with 5.11 fewer fetal losses compared to expected per 100 pregnancies for influenza vaccination during weeks 8-13 (Tables 1 and Figure 2(b)). The comparative control cohorts, corresponding to vaccination prior to pregnancy, also exhibited observed-to-expected patterns that were very different from COVID-19 vaccination during weeks 8-13, with women vaccinated prior to pregnancy exhibiting either slightly *lower-than-expected* or *according-to-expected* observed numbers (Figures 3 and S3).

**Figure 3.**
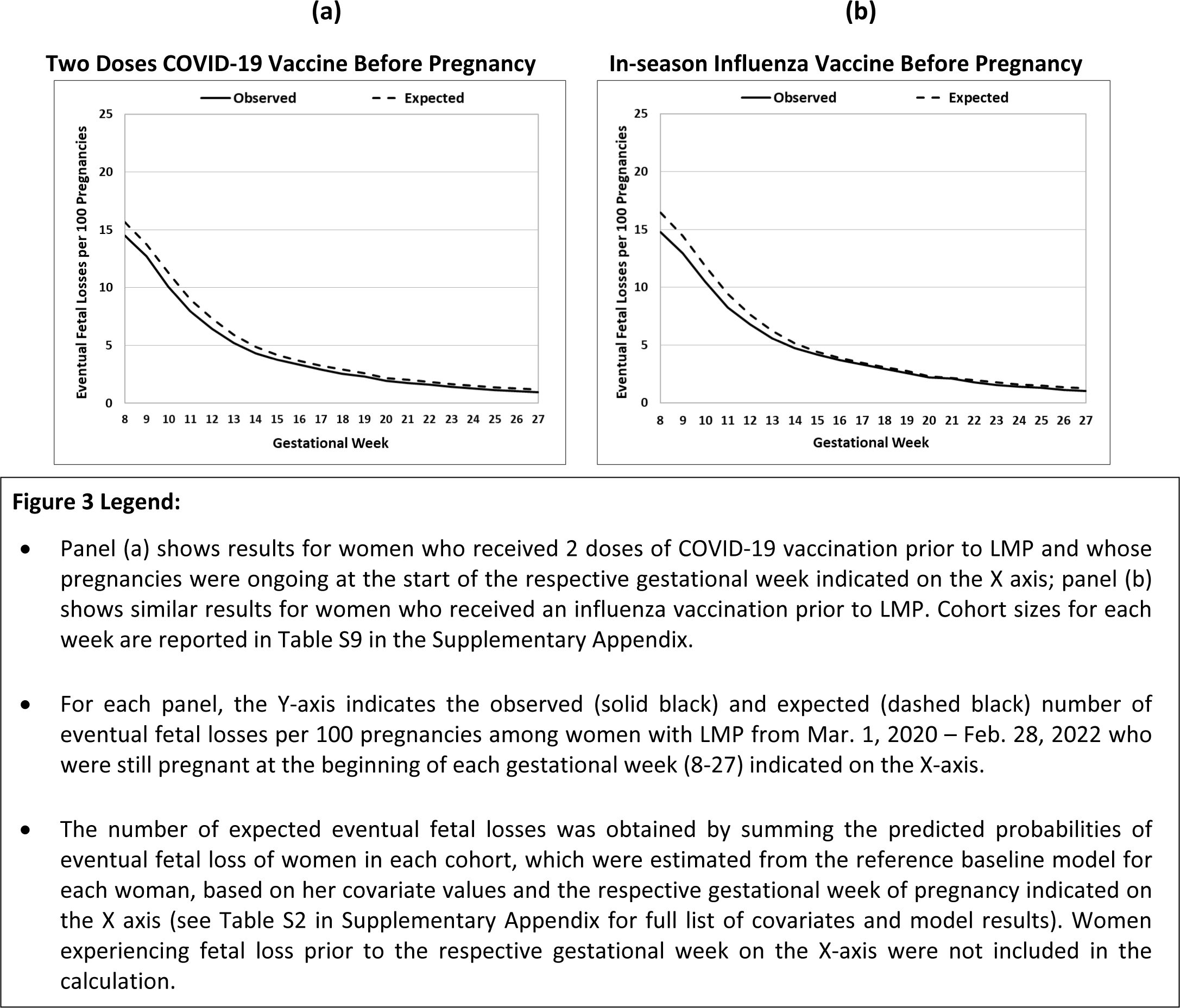
Observed-to-Expected Eventual Fetal Losses among Women Vaccinated for COVID-19 or Influenza Prior to Pregnancy.

There is a marked difference in observed-to-expected patterns of COVID-19 vaccinations between gestational weeks 8-13 and weeks 14-27 or prior to pregnancy. This difference, coupled with the *higher-than-expected* number of late fetal losses associated with vaccination during weeks 8-13 (including from week 25 onward), may be consistent with underlying biological mechanisms that depend on the timing of exposure during pregnancy. Indeed, exposure to teratogens, especially in early phases of pregnancy, is a known risk for adverse pregnancy outcomes and has been discussed in the context of vaccination, including influenza vaccines. (Bednarczyk and Adjaye-Gbewonyo 2012, Skowronski 2009) The critical period for the development of many congenital abnormalities overlaps with the early stages of pregnancy and specifically the 1^st^ trimester. (Czeizel 2008) Moreover, recent evidence suggests that transplacental transmission of COVID-19 mRNA vaccines occurs (Lin, et al. 2024, Chen, et al. 2025), and an in-vitro study shows suppression of embryo-fetal globin genes in human erythroleukemia K562 cells following exposure to the BNT162b2 vaccine. (Zurlo, et al. 2023) Additionally, mRNA COVID-19 vaccines are associated with thrombotic adverse events, which is another known inciting mechanism of spontaneous abortions. (Berild, et al. 2022)

Notably, the observed-to-expected differences for women receiving doses 1 and 2 during weeks 8-13 (87% of all women who received dose 1 continued to receive dose 2 during pregnancy) were 2-fold higher compared to those for women who received two doses prior to pregnancy and a single dose 3 during weeks 8-13 (3.85 vs. 1.9, respectively). This potential dose-response relationship may also be consistent with biological mechanisms, because the observed-to-expected difference increases with an additional dose received early in pregnancy.

It is plausible that the *lower-than-expected* observed numbers of fetal losses for both COVID-19 vaccination during gestational weeks 14-27 and influenza vaccination during weeks 8-27 were the result of healthy vaccinee bias, rather than a protective effect via vaccination. There is evidence from multiple countries, including Israel, of healthy vaccinee bias in studies on COVID-19 vaccination and hospitalization and death outcomes. (Høeg, Duriseti and Prasad 2023, Riedmann, et al. 2024, Xu, et al. 2021, Furst and Straka 2024) Healthy vaccinee bias, a type of healthy user bias, has also been highlighted in safety and efficacy studies of influenza vaccination, including during pregnancy. (Remschmidt, Wichmann and Harder 2015, Wolfe, et al. 2023, Dozelli 2018) Moreover, the observed-to-expected results for exposure to SARS-CoV-2 infections during pregnancy (Table S8) did not show any signal of significant increase in the number of fetal losses compared to expected even among unvaccinated women. Similarly, the infection rates during the 2020-2021 influenza season were significantly lower than previous years, including for example the 2018-2019 season. (Fratty, et al. 2022) Since the observed-to-expected differences for influenza vaccination during pregnancy in both periods were largely similar, it is unlikely that influenza vaccination reduced fetal losses significantly. While the *higher-than-expected* numbers of fetal losses associated with exposure to COVID-19 vaccination during gestational weeks 8-13 raises concern, the observed-to-expected analysis is not sufficient to establish a causal relationship.

Even if COVID-19 vaccination early in pregnancy truly increases the number of fetal losses above what is expected, any cumulative population-level impact would have likely been small and not easily detectable by traditional pharmacovigilance mechanisms. The reason for this is that only a relatively small number of women received vaccination during gestational weeks 8-13. In particular, out of the 20,383 women in this study who could have potentially received dose 1 during gestational weeks 8-27 based on their LMP, only 1,837, about 9%, received dose 1 during gestational weeks 8-13. Notably, these women seem to have had a lower *a priori* risk of fetal loss compared to the population. Moreover, 3.85 additional fetal losses per 100 pregnancies amounts to a total of 71 additional fetal losses above expected, which is less than a 0.35% of the 20,383 women. Such a signal could have been easily masked by random variability and temporal trends like the noted trend of decreasing rates of elective induced abortions in Israel, particularly during 2020-2022. (Israel Ministry of Health 2023) Indeed, as can be calculated from Figure 1, the overall percentage of pregnancies that resulted in fetal loss from gestational week 8 was 14% during 2020-2022, compared to 15.08% and 14.7% in 2018-2019 and 2016-2018, respectively.

This highlights the importance of developing robust pharmacovigilance methods, including prospective ones, to detect weak safety signals in sub-groups of the population. This is especially true for common outcomes such as miscarriage, premature birth and fetal abnormalities, which are unlikely to be reported as suspected adverse events unless there are unusual or pathognomonic characteristics, and where the window of teratogenicity may be narrow.

Much of the findings of this study, particularly the observed-to-expected analyses, cannot be directly compared to prior work, which focused almost entirely on risk-adjusting regression analyses that compare vaccinated to unvaccinated women during vaccination campaigns and mostly studied the impact of vaccination during later stages of pregnancy. (Rimmer, et al. 2023, Prasad, et al. 2022) However, such approaches are often challenged to obtain stable and similar cohorts, even when employing covariate balancing methods, such as inverse probability of treatment weighting (IPTW) and matching. (McCaffrey, et al. 2013, Austin 2015, Stuart 2010). Two studies on the association between COVID-19 vaccination during pregnancy and the incidence of SARS-CoV-2 infection using data from pregnancy registries in Israel illustrate these limitations. (Goldshtein, Nevo, et al. 2021, Dagan, et al. 2021) The studies were able to successfully match only 63% and 38% of the eligible vaccinated women, respectively. Moreover, 42% and 47% of the matched vaccinated-unvaccinated pairs, respectively, were censored when matched control women were vaccinated, leading to a short follow-up time. In contrast, the observed-to-expected analysis in this study allowed maximal matching of vaccinated women to historical (‘synthetic’) controls, longer follow up times and assessment of a range of vaccination exposures in terms of doses and timing.

Research from Scotland (Calvert, Carruthers, et al. 2022) is somewhat of an exception in that it matched vaccinated pregnant women to pregnant women with the same maternal age and gestational week from prior years. Although the authors did not report on any significant concerning findings, they did not consider fetal losses after week 20 and their analysis combined exposure starting at 6 weeks preconception with exposure through gestational week 20. Moreover, their matching algorithm was coarse and did not consider many important covariates with known impact on the risk of fetal loss, with the matched cohorts differing significantly. In contrast, the baseline reference model used in the current study relied on all pregnancies with LMP From March 2016 through February 2018 to establish individual adjusted risk scores that consider the specific covariates of the vaccinated women and cohort-specific expected fetal losses throughout the entire pregnancy.

A unique aspect of the current study compared to existing literature on the impact of COVID-19 vaccination during pregnancy was the use of multiple comparative controls. Comparing rates of the same adverse outcomes across different vaccines is commonly used in safety signal detection. (Vellozzi, et al. 2010) While women who were vaccinated for COVID-19 during pregnancy did not have identical characteristics to those who vaccinated for influenza during pregnancy, it is reasonable to assume that the two cohorts shared at least some unobserved confounding factors. Similarly, comparing different timing of vaccination has been previously used to assess potential safety risks of childhood vaccines. (DeStefano, et al. 2001, Velez, et al. 2023)

### Limitations

First, the observed-to-expected analysis is appropriate to detect potential safety signals but not to infer a causal relationship between vaccination and increased or decreased fetal loss rates or quantify the magnitude of such potential impact. Like any observational study, there are concerns regarding unobserved confounding covariates. However, if there were, such unobserved confounders would have had to be unique to women who received vaccination for COVID-19 but not for influenza. They also would have needed to be relevant to women who vaccinated during gestational weeks 8-13 but not during weeks 14-27 or prior to pregnancy. Second, this study did not include gestational weeks 1-7 and therefore could not assess the potential impact of vaccination during that period. Third, another general limitation of studies that are based on pregnancy registries stems from the fact that early fetal losses are often not documented, and more generally, the follow up may not be consistent across women. Indeed, there were pregnancies in the registry that were excluded because of lack of appropriate and timely follow-up, and while the analysis did not point to any obvious related biases, it cannot rule them out. Fourth, the Maccabi registry, like most pregnancy registries, did not provide sufficient information to distinguish between purely elective and medically-driven induced abortions. Thus, it is possible that some of the observed-to-expected differences are driven by behavioral mechanisms. For example, women who were vaccinated early in pregnancy may have been more likely to report fetal losses. Finally, this study is based only on data from Israel, and it would be important to conduct studies with similar data from other countries to see whether the results replicate.

### Conclusion

Overall, the findings in this paper provided concerning evidence of a *higher-than-expected* fetal loss rate associated with mRNA COVID-19 vaccine doses received during early pregnancy (gestational weeks 8-13). The safety signal should be further investigated by regulatory authorities as part of their risk assessment of vaccination during pregnancy with specific focus on the physiological effects in early pregnancy. There is also a need to conduct pathophysiological studies to better understand the potential biological mechanisms. Additionally, it would be insightful to assess the potential impact of non-mRNA COVID-19 vaccines. The findings also underscore the importance of conducting dedicated and statistically powered prospective clinical trials to study the impact of vaccination for COVID-19 and other pathogens during pregnancy to better inform recommendations to this vulnerable population.

## AUTHOR’S ROLES

Conceptualization: JG and RL; Acquisition of data: TP and SG; Data Analysis: JG and RL; Drafting: JG and RL; Interpretation of Results and Critical Review: JG, RL, TP, SG, TBH, JF, YS; Final Approval: JG, RL, TP, SG, TBH, JF, YS; Accountability: JG, RL, TP, SG, TBH, JF, YS

## Data Availability

Regulatory restrictions do not allow data sharing due to privacy concerns

## ACKNOWLEDGMENTS

The authors acknowledge helpful feedback and contributions to earlier versions of the paper from Jay Bhattacharya. Additional feedback was provided by Joseph Ladapo, Shoshy Altuvia, Efrat Schur, Alexandra Kalev, Norman Fenton, Clare Craig, Jessica Rose and Sam Saidi.

## Supplementary Appendix

**Table S1.**
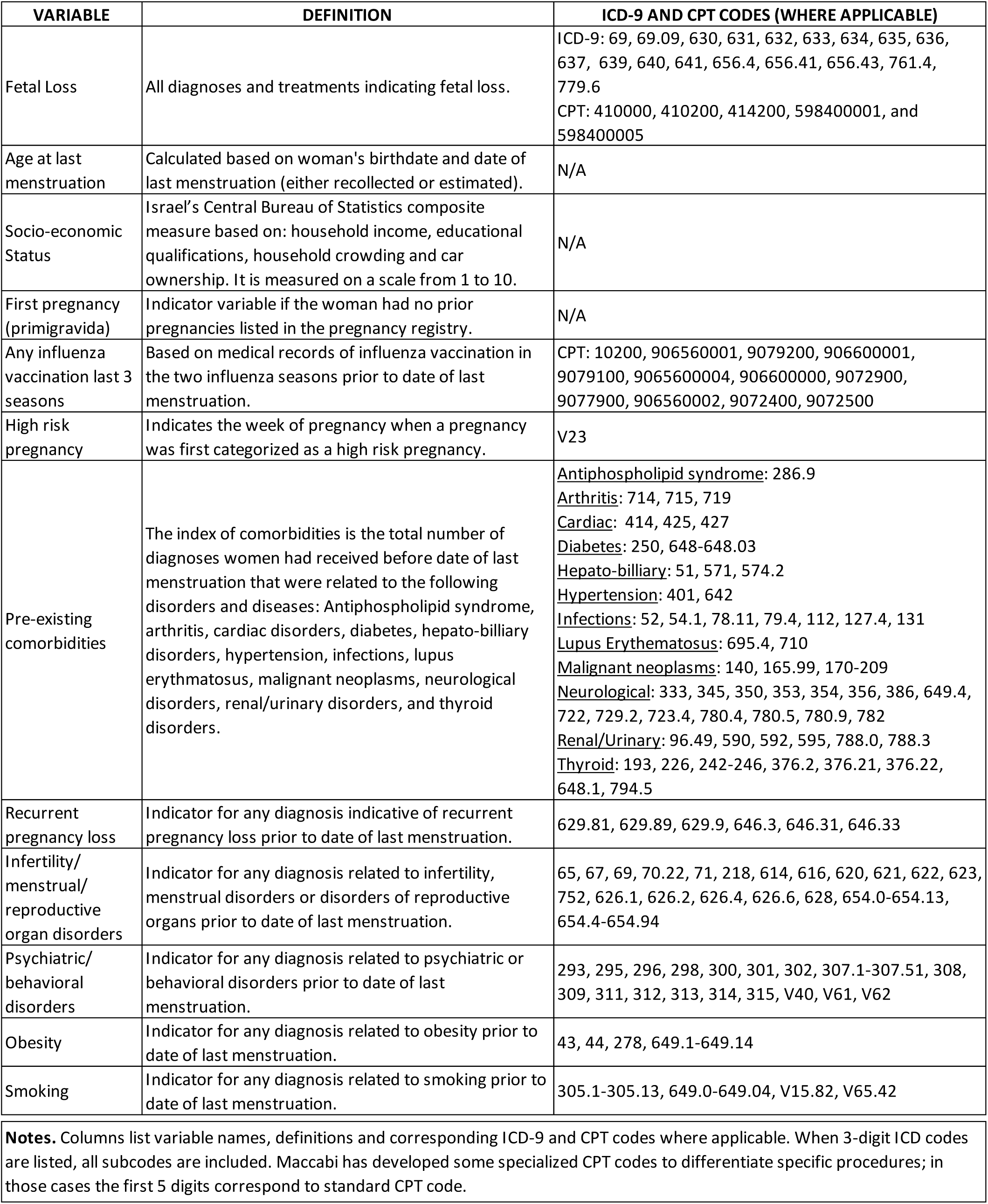
Dependent and Independent Variable Definitions for Baseline Regression Model.

| VARIABLE | DEFINITION | ICD-9 AND CPT CODES (WHERE APPLICABLE) |
| --- | --- | --- |
| Fetal Loss | All diagnoses and treatments indicating fetal loss. | ICD-9: 69, 69.09, 630, 631, 632, 633, 634, 635, 636, 637, 639, 640, 641, 656.4, 656.41, 656.43, 761.4, 779.6<br>CPT: 410000, 410200, 414200, 598400001, and 598400005 |
| Age at last menstruation | Calculated based on woman's birthdate and date of last menstruation (either recollected or estimated). | N/A |
| Socio-economic Status | Israel's Central Bureau of Statistics composite measure based on: household income, educational qualifications, household crowding and car ownership. It is measured on a scale from 1 to 10. | N/A |
| First pregnancy (primigravida) | Indicator variable if the woman had no prior pregnancies listed in the pregnancy registry. | N/A |
| Any influenza vaccination last 3 seasons | Based on medical records of influenza vaccination in the two influenza seasons prior to date of last menstruation. | CPT: 10200, 906560001, 9079200, 906600001, 9079100, 9065600004, 906600000, 9072900, 9077900, 906560002, 9072400, 9072500 |
| High risk pregnancy | Indicates the week of pregnancy when a pregnancy was first categorized as a high risk pregnancy. | V23 |
| Pre-existing comorbidities | The index of comorbidities is the total number of diagnoses women had received before date of last menstruation that were related to the following disorders and diseases: Antiphospholipid syndrome, arthritis, cardiac disorders, diabetes, hepato-biliary disorders, hypertension, infections, lupus erythematosus, malignant neoplasms, neurological disorders, renal/urinary disorders, and thyroid disorders. | <u>Antiphospholipid syndrome</u>: 286.9<br><u>Arthritis</u>: 714, 715, 719<br><u>Cardiac</u>: 414, 425, 427<br><u>Diabetes</u>: 250, 648-648.03<br><u>Hepato-biliary</u>: 51, 571, 574.2<br><u>Hypertension</u>: 401, 642<br><u>Infections</u>: 52, 54.1, 78.11, 79.4, 112, 127.4, 131<br><u>Lupus Erythematosus</u>: 695.4, 710<br><u>Malignant neoplasms</u>: 140, 165.99, 170-209<br><u>Neurological</u>: 333, 345, 350, 353, 354, 356, 386, 649.4, 722, 729.2, 723.4, 780.4, 780.5, 780.9, 782<br><u>Renal/Urinary</u>: 96.49, 590, 592, 595, 788.0, 788.3<br><u>Thyroid</u>: 193, 226, 242-246, 376.2, 376.21, 376.22, 648.1, 794.5 |
| Recurrent pregnancy loss | Indicator for any diagnosis indicative of recurrent pregnancy loss prior to date of last menstruation. | 629.81, 629.89, 629.9, 646.3, 646.31, 646.33 |
| Infertility/ menstrual/ reproductive organ disorders | Indicator for any diagnosis related to infertility, menstrual disorders or disorders of reproductive organs prior to date of last menstruation. | 65, 67, 69, 70.22, 71, 218, 614, 616, 620, 621, 622, 623, 752, 626.1, 626.2, 626.4, 626.6, 628, 654.0-654.13, 654.4-654.94 |
| Psychiatric/ behavioral disorders | Indicator for any diagnosis related to psychiatric or behavioral disorders prior to date of last menstruation. | 293, 295, 296, 298, 300, 301, 302, 307.1-307.51, 308, 309, 311, 312, 313, 314, 315, V40, V61, V62 |
| Obesity | Indicator for any diagnosis related to obesity prior to date of last menstruation. | 43, 44, 278, 649.1-649.14 |
| Smoking | Indicator for any diagnosis related to smoking prior to date of last menstruation. | 305.1-305.13, 649.0-649.04, V15.82, V65.42 |
| **Notes.** Columns list variable names, definitions and corresponding ICD-9 and CPT codes where applicable. When 3-digit ICD codes are listed, all subcodes are included. Maccabi has developed some specialized CPT codes to differentiate specific procedures; in those cases the first 5 digits correspond to standard CPT code. |  |  |

### Section S1. Description of Baseline Reference Regression Model in the *Reference* period

The baseline reference model used to estimate the time-dependent conditional risk probability of eventual fetal loss is a pooled logistic regression that includes an observation for every woman and week of pregnancy (R. D’Agostino, et al. 1990). Since this study is concerned with an uncensored binary clinical outcome (fetal loss or live birth) and not with time-to-event, a logistic regression model is a natural modeling choice (Hu and Tong 2021). The model is trained on all pregnancies within the *Reference* period (March 1, 2016 – February 28, 2018) that survived to the start of gestational week 8, had a documented birth outcome and include complete information on the covariates. Pregnancies that ended prior to the start of week 8 were excluded, because a significant number of them end without appropriate and consistent documentation, which is a known source of bias in estimating miscarriage rates (Howards and Hertz-Picciotto 2007). For each pregnancy, there is an observation for each gestational week 8-27 (i.e., until end of the 2^nd^ trimester), as long as the pregnancy was ongoing at the start of that week. The unit of analysis is pregnancy-week. The dependent (outcome) variable for each observation equals 1 if fetal loss eventually occurred at any point from the start of the respective gestational week, and 0 otherwise. The independent variables of the baseline reference model include the gestational week, calendar month of LMP, maternal age, socioeconomic status, primigravida status, an aggregated count (0, 1-3 and 4 or more) of medical comorbidities with potential impact on the study’s outcome, social sector, district of residency, health-related behaviors (e.g., smoking and influenza vaccination in the two seasons prior to the index pregnancy), and an indicator for whether the pregnancy was considered high-risk that equals 1 starting from the week in which the pregnancy was considered high risk. The comorbidities included in the model were selected if they appeared in significantly higher frequency among women who experienced fetal loss compared to women who had a live birth. See Table S1 for covariate definitions. Confidence intervals were corrected to account for multiple observations from a single pregnancy across weeks using the Liang-Zeger adjustment (Liang and Zeger 1986, Abadie, Athey and Imbens 2023).

**Table S2.** Baseline Pooled Logistic Regression of Eventual Fetal Loss in the *Reference* Period (Mar. 1, 2016 – Feb. 28, 2018)

| **Coefficients** | **Odds Ratio** | **95% Confidence Interval** |
| --- | --- | --- |
| Age at last menstruation | 1.084 | 1.079 - 1.090 |
| Socio-economic status | 0.951 | 0.938 - 0.964 |
| First pregnancy (primigravida) indicator | 2.088 | 1.969 - 2.214 |
| Any influenza vaccination last 3 seasons | 1.078 | 1.011 - 1.150 |
| High risk pregnancy (first two trimesters) | 2.212 | 1.784 - 2.743 |
| **Diagnoses prior to pregnancy** |  |  |
| 0 pre-existing comorbidities (reference category) | 1.000 | 1.000 - 1.000 |
| 1 - 3 pre-existing comorbidities | 1.144 | 1.076 - 1.216 |
| 4+ pre-existing comorbidities | 1.223 | 1.070 - 1.397 |
| Recurrent pregnancy loss | 1.718 | 1.442 - 2.046 |
| Infertility/menstrual/reproductive organ disorders | 1.207 | 1.137 - 1.281 |
| Psychiatric/behavioral disorders | 1.118 | 1.055 - 1.184 |
| Obesity | 1.056 | 0.987 - 1.129 |
| Smoking | 1.053 | 0.980 - 1.132 |
| **District of residency** |  |  |
| North | 0.855 | 0.780 - 0.937 |
| Sharon | 1.017 | 0.938 - 1.102 |
| South | 1.087 | 0.997 - 1.185 |
| Jerusalem | 0.916 | 0.847 - 0.990 |
| Center (reference category) | 1.000 | 1.000 - 1.000 |
| **Ethno-religious sector** |  |  |
| Arab | 0.866 | 0.757 - 0.991 |
| Orthodox | 0.854 | 0.753 - 0.969 |
| Ultra Orthodox | 0.778 | 0.699 - 0.866 |
| Other (reference category) | 1.000 | 1.000 - 1.000 |

**Table S2 (cont'd). Baseline Pooled Logistic Regression of Eventual Fetal Loss in the *Reference* Period**
| **Coefficients** | **Odds Ratio** | **95% Confidence Interval** |
| --- | --- | --- |
| **Gestational week** |  |  |
| 8 (reference category) | 1.000 | 1.000 - 1.000 |
| 9 | 0.856 | 0.850 - 0.862 |
| 10 | 0.686 | 0.678 - 0.694 |
| 11 | 0.533 | 0.524 - 0.541 |
| 12 | 0.423 | 0.414 - 0.431 |
| 13 | 0.337 | 0.329 - 0.345 |
| 14 | 0.275 | 0.267 - 0.283 |
| 15 | 0.232 | 0.225 - 0.240 |
| 16 | 0.205 | 0.198 - 0.212 |
| 17 | 0.181 | 0.174 - 0.187 |
| 18 | 0.160 | 0.154 - 0.166 |
| 19 | 0.143 | 0.137 - 0.149 |
| 20 | 0.117 | 0.112 - 0.123 |
| 21 | 0.111 | 0.105 - 0.116 |
| 22 | 0.099 | 0.094 - 0.105 |
| 23 | 0.089 | 0.084 - 0.094 |
| 24 | 0.081 | 0.077 - 0.086 |
| 25 | 0.075 | 0.070 - 0.080 |
| 26 | 0.069 | 0.064 - 0.073 |
| 27 | 0.063 | 0.059 - 0.067 |
| **Calendar month** |  |  |
| January | 1.014 | 0.896 - 1.149 |
| February | 0.981 | 0.864 - 1.115 |
| March (reference category) | 1.000 | 1.000 - 1.000 |
| April | 1.001 | 0.882 - 1.135 |
| May | 1.053 | 0.930 - 1.193 |
| June | 1.087 | 0.959 - 1.231 |
| July | 1.080 | 0.953 - 1.224 |
| August | 1.105 | 0.976 - 1.252 |
| September | 1.061 | 0.936 - 1.202 |
| October | 0.982 | 0.867 - 1.111 |
| November | 1.101 | 0.973 - 1.245 |
| December | 1.025 | 0.907 - 1.159 |
| **Constant** | 0.011 | 0.009 - 0.013 |
| N =1,561,906 |  |  |

### Section S2. Formula for Calculating Expected Values and Confidence Intervals for Eventual Fetal Loss Rate

The baseline reference regression model estimates are used to obtain the risk probability of eventual fetal loss for every gestational week for each pregnancy included in the cohort. The probability is estimated prospectively considering the gestational week of the pregnancy, which is typically the week of vaccination or a later week (assuming the pregnancy was ongoing at the start of that week), as well as the calendar month of the LMP and other covariates in the baseline model. To obtain a cohort-specific expected number of fetal losses, the individual risk probabilities are summed. That is, if there are N pregnancies in the cohort, the cohort-specific expected rate is 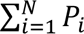, where *P_i_* denotes the predicted risk probability of eventual fetal loss of pregnancy *i*. The expected number of fetal losses is normalized to number of fetal losses per 100 pregnancies, i.e., 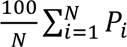.

To calculate the CIs, we assume that the cohort-specific number of fetal losses is the sum of independent Bernoulli variables, each corresponding to a specific pregnancy, with probability *P_i_* of being equal to 1 (i.e., having a fetal loss). It is sufficient to calculate a CI for 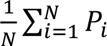. From the assumption that the number of fetal losses is the sum of independent Bernoulli variables, it follows that the variance is equal to 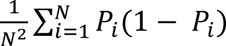, and using Lyapunov’s Central Limit Theorem the CI is:

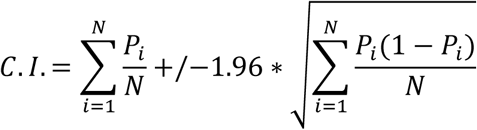

**Figure S1.**
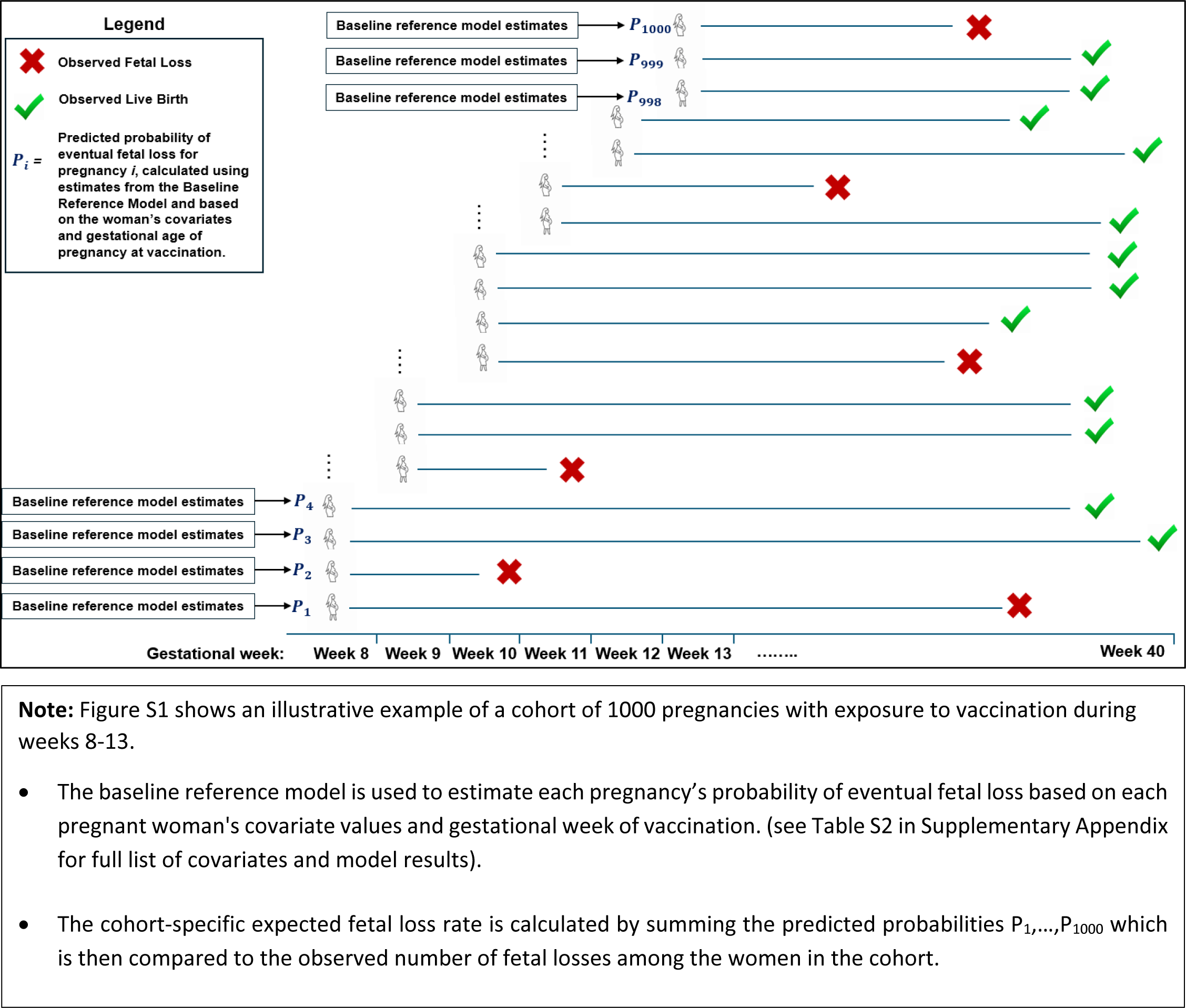
Diagram Illustrating Methodological Approach Applied to Cohort of Women Vaccinated in Gestational Weeks 8-13.

**Table S3.** Characteristics of Pregnant Women in *Reference*, *Validation* and *COVID-19* Periods.

| Characteristics | Reference Period |  | Validation Period |  | COVID-19 Period |  |
| --- | --- | --- | --- | --- | --- | --- |
|  | Mean | 95% CI | Mean | 95% CI | Mean | 95% CI |
| Age at last menstruation | **30.31** | 30.27 - 30.35 | **30.23** | 30.17 - 30.28 | **30.40** | 30.19 - 30.27 |
| Socio-economic status | **5.71** | 5.69 - 5.73 | **5.55** | 5.52 - 5.57 | **5.52** | 5.46 - 5.50 |
|  | **%** |  | **%** |  | **%** |  |
| First pregnancy (primigravida) indicator | **40.43** | 40.08 - 40.78 | **38.55** | 38.09 - 39.01 | **41.40** | 41.07 - 41.74 |
| Any influenza vaccination last 3 seasons | **21.67** | 21.38 - 21.96 | **23.85** | 23.45 - 24.26 | **27.55** | 27.25 - 27.85 |
| High risk pregnancy (first two trimesters) | **0.14** | 0.11 - 0.17 | **0.12** | 0.09 - 0.16 | **0.13** | 0.11 - 0.16 |
| **Diagnoses prior to pregnancy** |  |  |  |  |  |  |
| 1 - 3 pre-existing comorbidities | **55.98** | 55.63 - 56.33 | **57.33** | 56.86 - 57.79 | **59.15** | 58.81 - 59.48 |
| 4+ pre-existing comorbidities | **3.46** | 3.33 - 3.59 | **3.44** | 3.26 - 3.61 | **3.41** | 3.28 - 3.53 |
| Recurrent pregnancy loss | **1.19** | 1.11 - 1.27 | **1.52** | 1.40 - 1.63 | **1.39** | 1.31 - 1.47 |
| Infertility/menstrual/reproductive organ disorders | **51.95** | 51.60 - 52.31 | **52.95** | 52.48 - 53.43 | **53.04** | 52.70 - 53.38 |
| Psychiatric/behavioral disorders | **30.22** | 29.90 - 30.55 | **31.94** | 31.49 - 32.38 | **34.46** | 34.14 - 34.78 |
| Obesity | **17.51** | 17.24 - 17.78 | **18.69** | 18.32 - 19.06 | **19.40** | 19.13 - 19.67 |
| Smoking | **14.06** | 13.81 - 14.30 | **13.51** | 13.19 - 13.84 | **12.68** | 12.45 - 12.90 |
| **District of residency** |  |  |  |  |  |  |
| North | **15.18** | 14.93 - 15.44 | **14.87** | 14.53 - 15.20 | **14.51** | 14.28 - 14.75 |
| Sharon | **20.59** | 20.30 - 20.87 | **20.16** | 19.78 - 20.54 | **19.23** | 18.96 - 19.49 |
| South | **16.88** | 16.61 - 17.14 | **16.43** | 16.08 - 16.78 | **16.16** | 15.91 - 16.41 |
| Jerusalem | **24.66** | 24.35 - 24.96 | **24.77** | 24.36 - 25.18 | **25.70** | 25.40 - 26.00 |
| Center | **22.70** | 22.40 - 22.99 | **23.76** | 23.36 - 24.17 | **24.40** | 24.11 - 24.69 |
| **Ethno-religious sector** |  |  |  |  |  |  |
| Arab | **6.06** | 5.89 - 6.23 | **6.40** | 6.17 - 6.63 | **6.74** | 6.57 - 6.91 |
| Orthodox | **5.16** | 5.00 - 5.31 | **5.29** | 5.08 - 5.50 | **5.46** | 5.31 - 5.61 |
| Ultra Orthodox | **16.99** | 16.72 - 17.25 | **19.19** | 18.82 - 19.56 | **19.65** | 19.38 - 19.92 |
| Other | **71.80** | 71.48 - 72.12 | **69.12** | 68.68 - 69.56 | **68.15** | 67.84 - 68.47 |

**Note:** The *Reference* period includes women with LMP from Mar. 1, 2016 – Feb. 28, 2018; the *Validation* period includes women with LMP from Mar. 1, 2018 – Feb. 28, 2019; and the *COVID-19* period includes women with LMP from Mar. 1, 2020 – Feb. 28, 2022.

**Table S4.** Observed-to-Expected Eventual Fetal Losses among Women Vaccinated for COVID-19 with Dose 2 in Gestational Weeks 8-13 and 14-27.

| Cohort | Observed<br>Fetal Losses<br>per 100<br>Pregnancies<br>(n) | Expected<br>Fetal Losses<br>per 100<br>Pregnancies<sup>a</sup><br>(n) | Observed-to-Expected<br>Difference<br>per 100<br>Pregnancies<br>(n) | 95% C.I.<br>Observed-to-Expected<br>Difference<br>per 100<br>Pregnancies | Cohort<br>Size<br>N |
| --- | --- | --- | --- | --- | --- |
| ***Dose 2*<sup>b</sup>** |  |  |  |  |  |
| **During weeks 8-13** | **14.11**<br>(187) | **10.51**<br>(139) | **3.61**<br>(48) | **[1.99 - 5.22]** | **1 325** |
| **During weeks 14-27** | **1.95**<br>(200) | **2.25**<br>(231) | **-0.30**<br>(-31) | **[-0.59 - -0.02]** | **10 266** |

**Notes:** Table S4 shows observed-to-expected fetal losses (per 100 pregnancies) among pregnant women with LMP from Mar. 1, 2020 – Feb. 28, 2022 vaccinated for COVID-19 with dose 2. Numbers (n) in parentheses report the corresponding total number of (observed/expected) fetal losses in the cohort.
a. The number of expected fetal losses was obtained by summing the predicted probabilities of eventual fetal loss of women vaccinated in each gestational week, which were estimated for each woman from the baseline reference model, based on her covariate values and gestational week of vaccination (see Table S2 in Supplementary Appendix for full list of covariates and model results).
b. Of women vaccinated with dose 2 during weeks 8-13 and weeks 14-27, 97% and 96% received BNT162b2.

**Figure S2.**
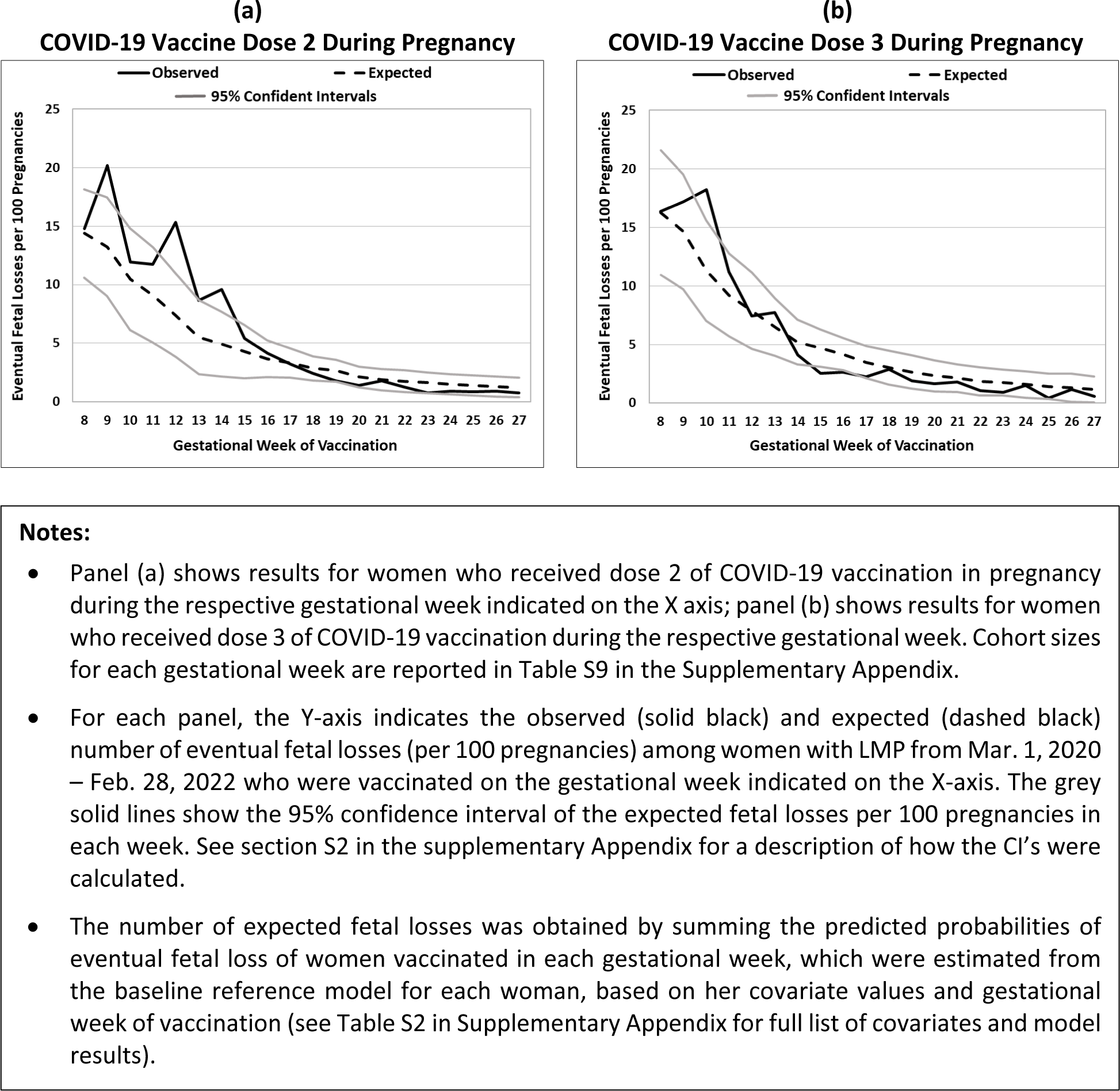
Observed-to-Expected Eventual Fetal Losses by Gestational Week of Vaccination among Women Vaccinated for COVID-19 with Dose 2 or Dose 3.

**Figure S3.**
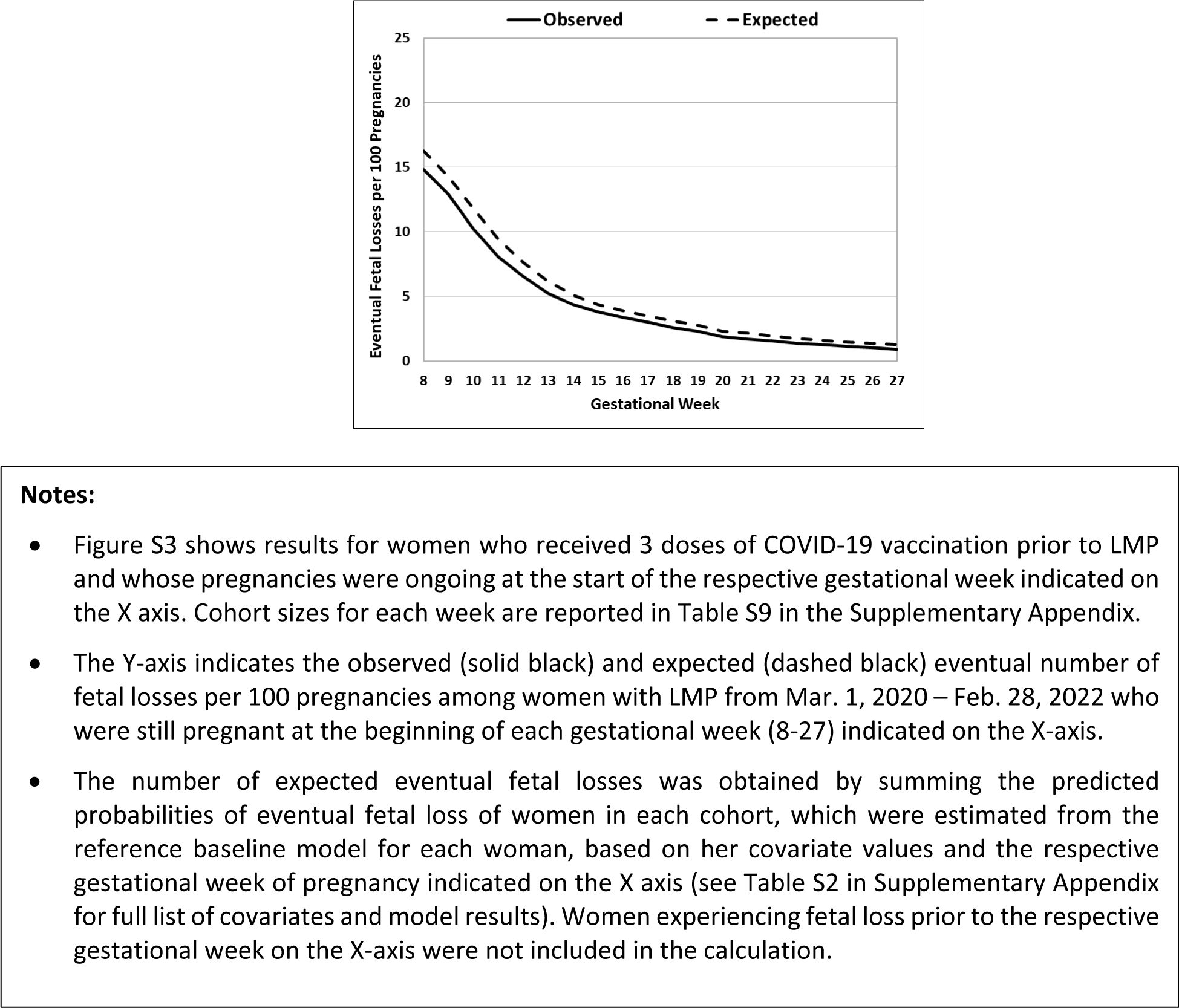
Observed-to-Expected Eventual Fetal Losses by Gestational Week among Women Vaccinated for COVID-19 with Dose 3 Prior to Pregnancy.

**Table S5.**
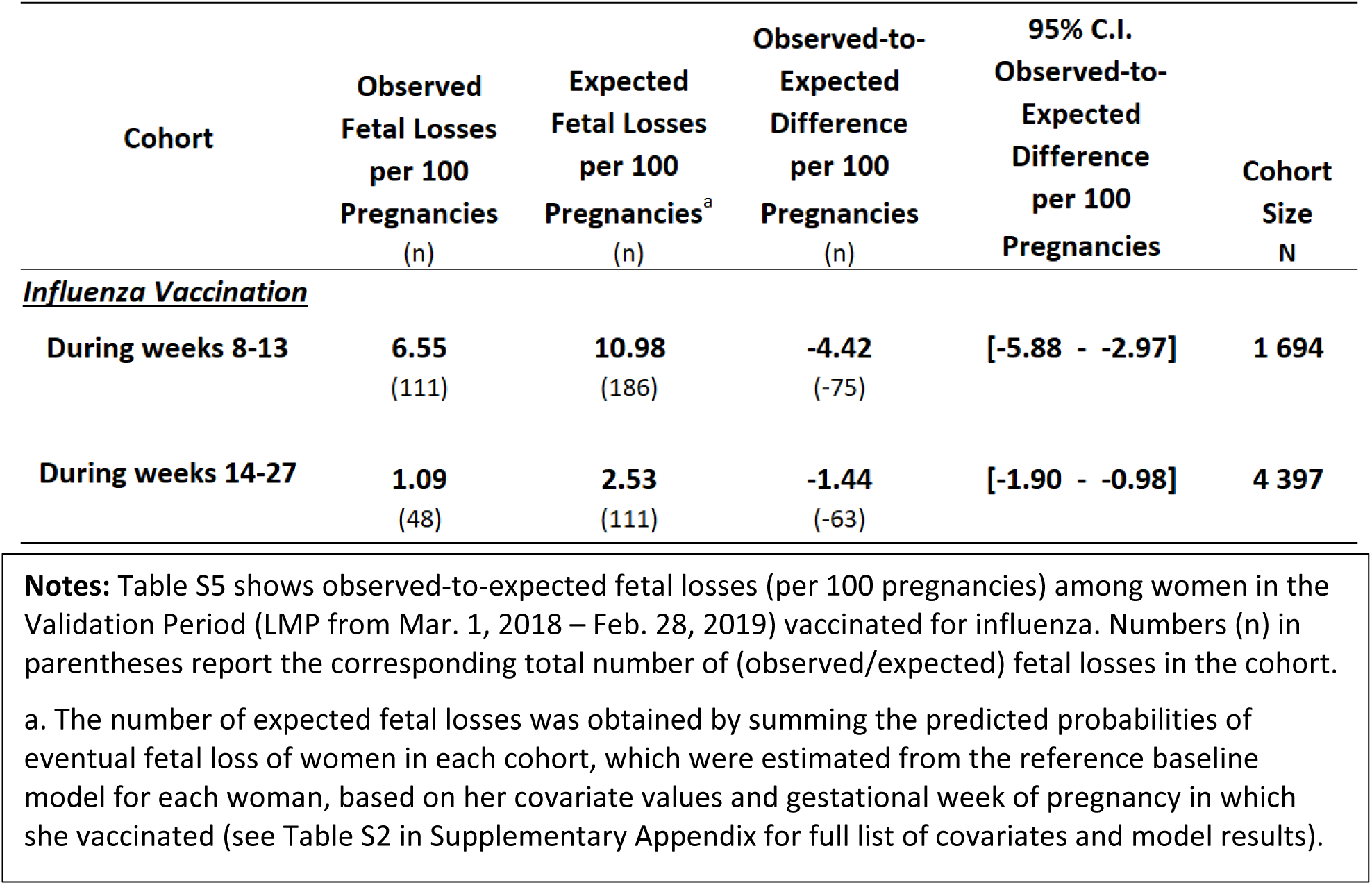
Observed-to-Expected Eventual Fetal Losses among Women Vaccinated for Influenza in Gestational Weeks 8-13 and 14-27 in the *Validation* Period (LMP from Mar. 1, 2018 – Feb. 28, 2019)

| Cohort | Observed<br>Fetal Losses<br>per 100<br>Pregnancies<br>(n) | Expected<br>Fetal Losses<br>per 100<br>Pregnancies<sup>a</sup><br>(n) | Observed-to-Expected<br>Difference<br>per 100<br>Pregnancies<br>(n) | 95% C.I.<br>Observed-to-Expected<br>Difference<br>per 100<br>Pregnancies | Cohort<br>Size<br>N |
| --- | --- | --- | --- | --- | --- |
| **<u>Influenza Vaccination</u>** |  |  |  |  |  |
| **During weeks 8-13** | **6.55**<br>(111) | **10.98**<br>(186) | **-4.42**<br>(-75) | **[-5.88 - -2.97]** | **1 694** |
| **During weeks 14-27** | **1.09**<br>(48) | **2.53**<br>(111) | **-1.44**<br>(-63) | **[-1.90 - -0.98]** | **4 397** |

**Notes:** Table S5 shows observed-to-expected fetal losses (per 100 pregnancies) among women in the Validation Period (LMP from Mar. 1, 2018 – Feb. 28, 2019) vaccinated for influenza. Numbers (n) in parentheses report the corresponding total number of (observed/expected) fetal losses in the cohort.
a. The number of expected fetal losses was obtained by summing the predicted probabilities of eventual fetal loss of women in each cohort, which were estimated from the reference baseline model for each woman, based on her covariate values and gestational week of pregnancy in which she vaccinated (see Table S2 in Supplementary Appendix for full list of covariates and model results).

**Figure S4.**
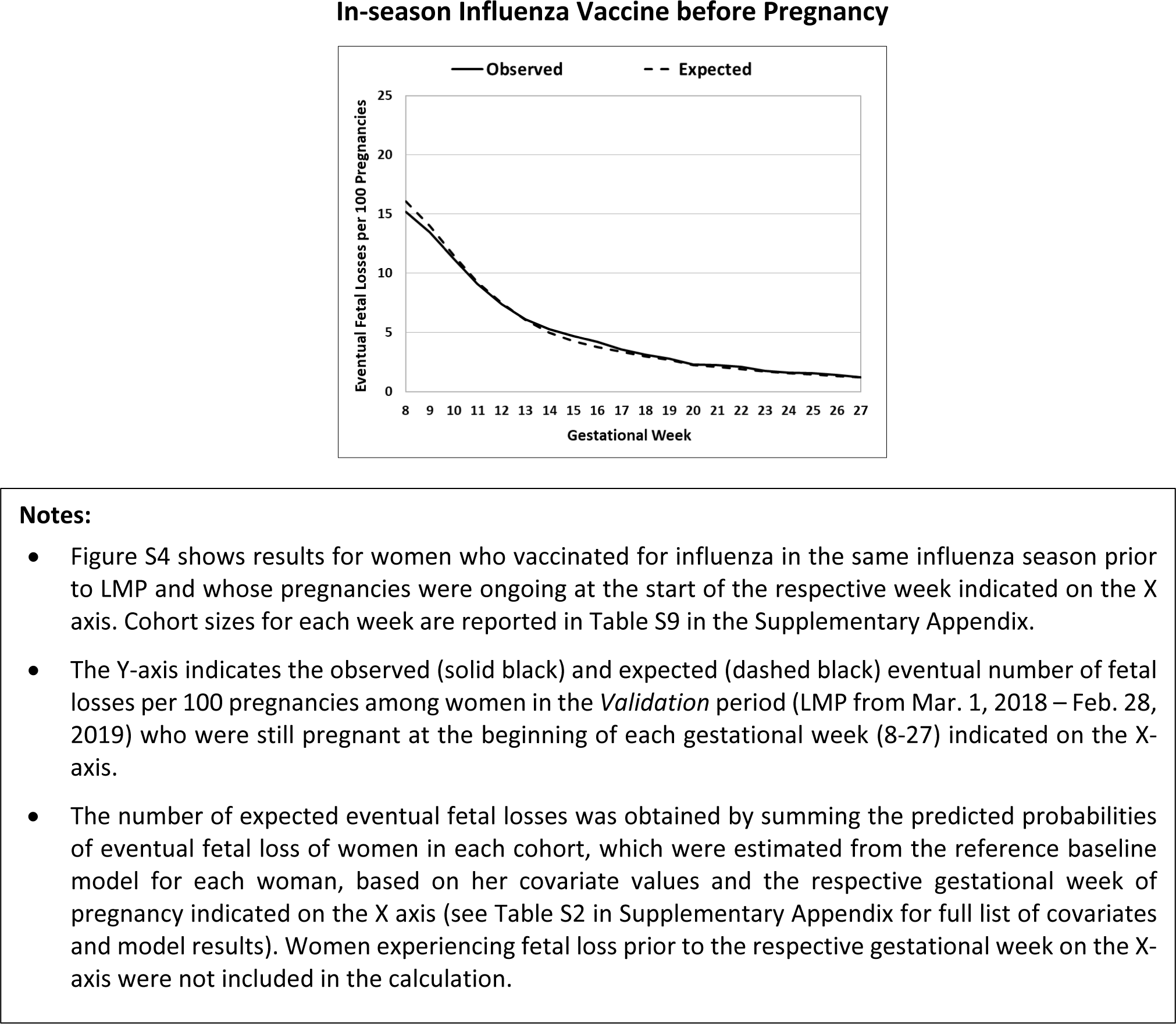
Observed-to-Expected Eventual Fetal Losses by Gestational Week among Women Vaccinated for Influenza Prior to Pregnancy with LMP in the *Validation* Period (LMP from Mar. 1, 2018 – Feb. 28, 2019)

**Table S6.** Observed-to-Expected Eventual Fetal Loss Ratios among All Cohorts of Women Vaccinated in Weeks 8-13 and 14-27.

| Cohort | Ratio of<br>Observed-to-Expected<br>per 100 Pregnancies<sup>a</sup> | 95% C.I. of Ratio of<br>Observed-to-Expected<br>per 100 Pregnancies | Cohort<br>Size<br>N |
| --- | --- | --- | --- |
| ***COVID-19 Dose 1<sup>b</sup>*** |  |  |  |
| During weeks 8-13 | 1.42 | [1.24 - 1.65] | 1 837 |
| During weeks 14-27 | 0.69 | [0.62 - 0.77] | 12 274 |
| ***COVID-19 Dose 2<sup>c</sup>*** |  |  |  |
| During weeks 8-13 | 1.34 | [1.16 - 1.59] | 1 325 |
| During weeks 14-27 | 0.87 | [0.77 - 0.99] | 10 266 |
| ***COVID-19 Dose 3<sup>b</sup>*** |  |  |  |
| During weeks 8-13 | 1.19 | [1.03 - 1.40] | 1 454 |
| During weeks 14-27 | 0.69 | [0.61 - 0.80] | 7 546 |
| ***Influenza Vaccine<sup>b</sup>*** |  |  |  |
| During weeks 8-13 | 0.55 | [0.50 - 0.60] | 4 048 |
| During weeks 14-27 | 0.50 | [0.45 - 0.57] | 9 586 |
| ***LATE FETAL LOSSES<sup>d</sup>*** |  |  |  |
| ***COVID-19 Dose 1*** |  |  |  |
| From week 14 | 1.64 | [1.36 - 2.07] | 1 733 |
| From week 20 | 2.16 | [1.63 - 3.19] | 1 674 |
| From week 25 | 2.22 | [1.58 - 3.75] | 1 646 |
| ***COVID-19 Dose 3*** |  |  |  |
| From week 14 | 1.53 | [1.25 - 1.97] | 1 389 |
| From week 20 | 1.66 | [1.23 - 2.54] | 1 330 |
| From week 25 | 1.64 | [1.14 - 2.94] | 1 311 |
| ***Influenza Vaccine*** |  |  |  |
| From week 14 | 0.57 | [0.50 - 0.66] | 3 918 |
| From week 20 | 0.06 | [0.51 - 0.77] | 3 856 |
| From week 25 | 0.05 | [0.40 - 0.66] | 3 828 |

Note: Table S6 shows the ratios of the number of observed-to-expected eventual fetal losses per 100 pregnancies that were included in this study.
a. The number of expected fetal losses was obtained by summing the predicted probabilities of eventual fetal loss of women in each cohort, which were estimated from the reference baseline model for each woman, based on her covariate values and gestational week of pregnancy in which she vaccinated (see Table S2 in Supplementary Appendix for full list of covariates and model results).
b. Based on results presented in Table 1.
c. Based on results presented in TableS4.
d. Based on results presented in Table 2.

**Table S7.**
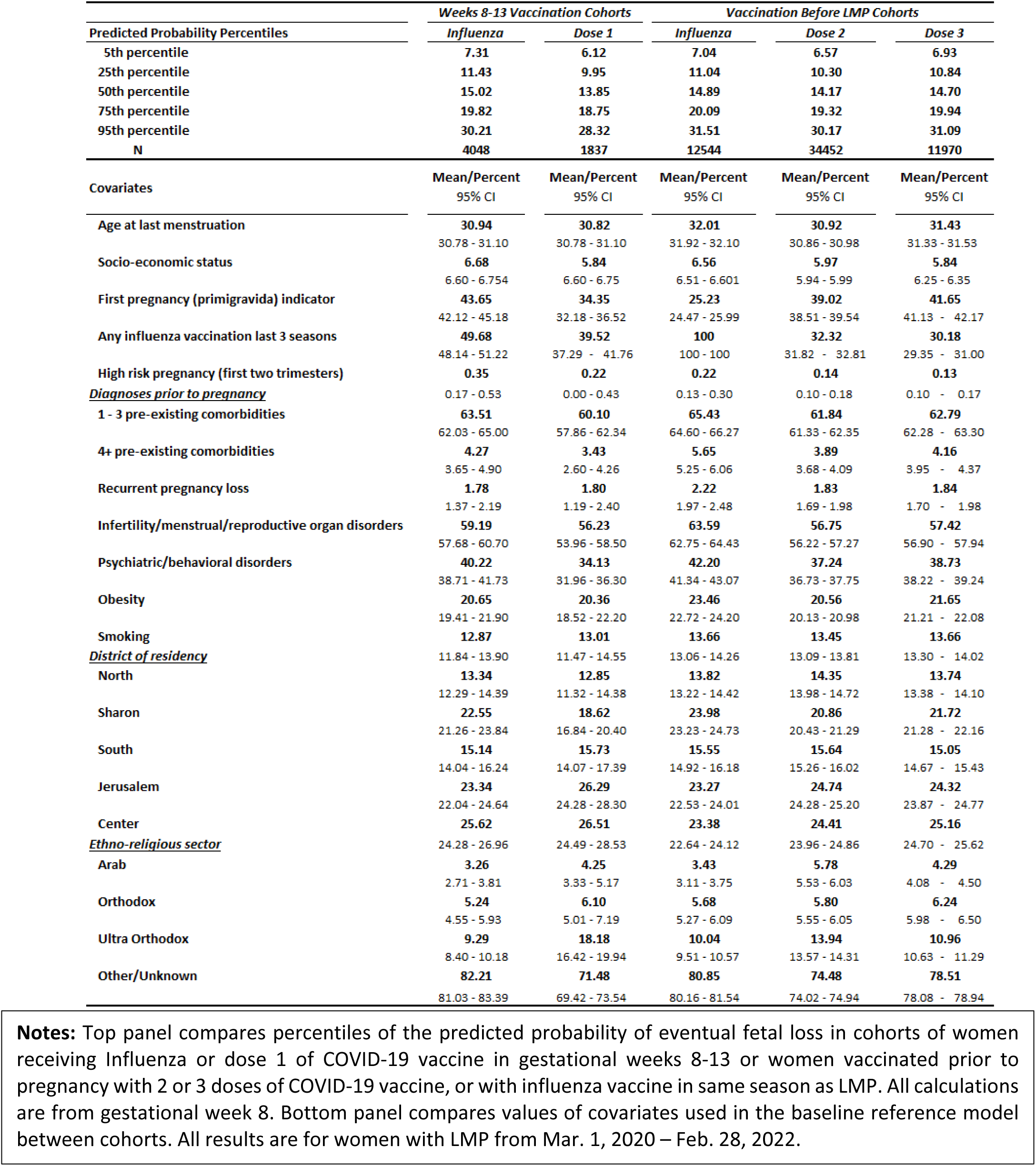
Percentile Distribution of Predicted Probability of Eventual Fetal Loss and Mean Values of Covariates for Women Vaccinated for Influenza and *COVID-19* with Dose 1 in weeks 8-13 and Vaccination Before LMP in the *COVID-19* Period

**Table S8.**
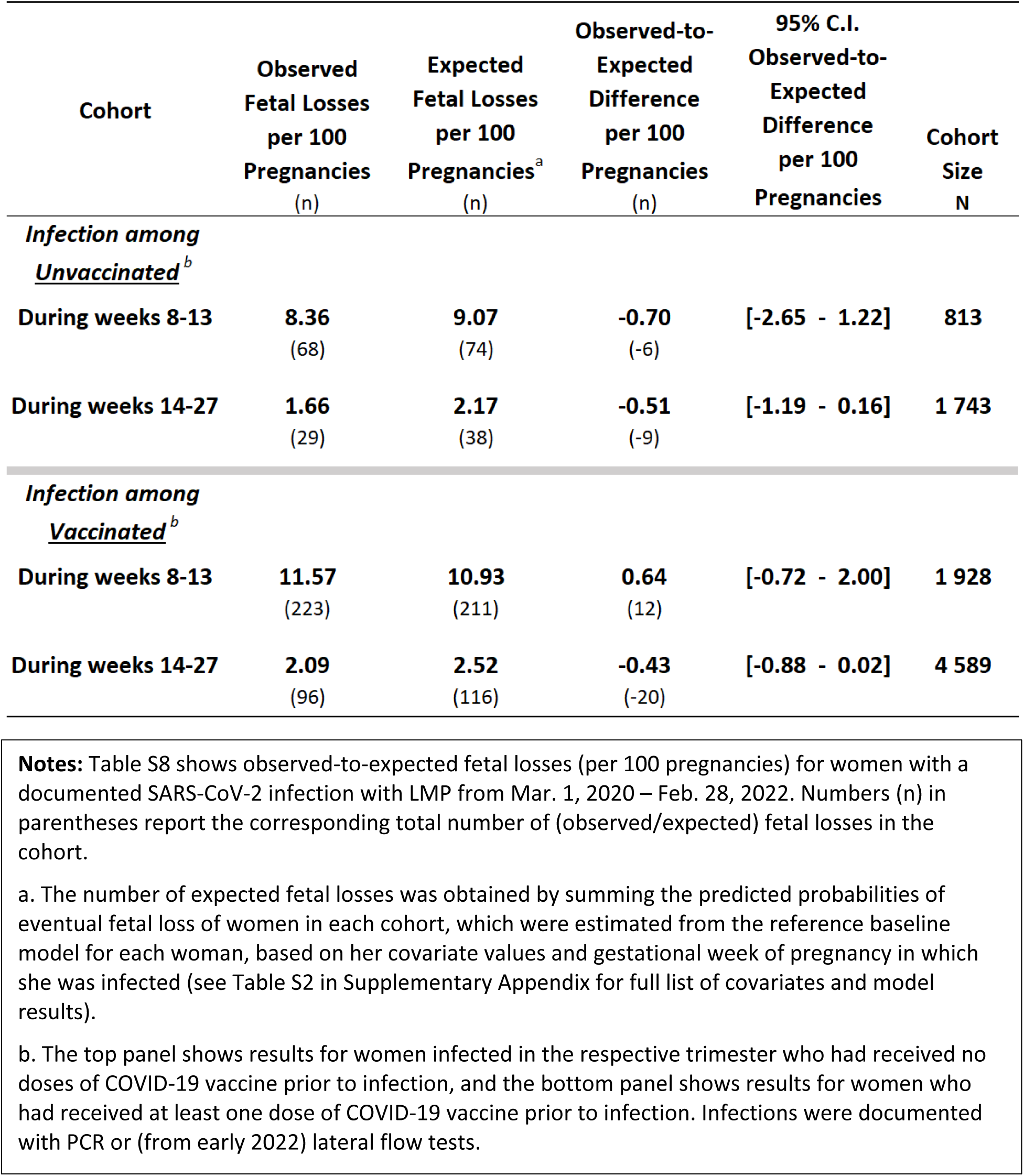
Observed-to-Expected Eventual Fetal Losses among Women with SARS-CoV-2 Infections in Gestational Weeks 8-13 and 14-27 by Vaccination Status.

| Cohort | Observed<br>Fetal Losses<br>per 100<br>Pregnancies<br>(n) | Expected<br>Fetal Losses<br>per 100<br>Pregnancies<sup>a</sup><br>(n) | Observed-to-Expected<br>Difference<br>per 100<br>Pregnancies<br>(n) | 95% C.I.<br>Observed-to-Expected<br>Difference<br>per 100<br>Pregnancies | Cohort<br>Size<br>N |
| --- | --- | --- | --- | --- | --- |
| ***Infection among***<br>***Unvaccinated<sup>b</sup>*** |  |  |  |  |  |
| **During weeks 8-13** | **8.36**<br>(68) | **9.07**<br>(74) | **-0.70**<br>(-6) | **[-2.65 - 1.22]** | **813** |
| **During weeks 14-27** | **1.66**<br>(29) | **2.17**<br>(38) | **-0.51**<br>(-9) | **[-1.19 - 0.16]** | **1 743** |
| ***Infection among***<br>***Vaccinated<sup>b</sup>*** |  |  |  |  |  |
| **During weeks 8-13** | **11.57**<br>(223) | **10.93**<br>(211) | **0.64**<br>(12) | **[-0.72 - 2.00]** | **1 928** |
| **During weeks 14-27** | **2.09**<br>(96) | **2.52**<br>(116) | **-0.43**<br>(-20) | **[-0.88 - 0.02]** | **4 589** |

**Notes:** Table S8 shows observed-to-expected fetal losses (per 100 pregnancies) for women with a documented SARS-CoV-2 infection with LMP from Mar. 1, 2020 – Feb. 28, 2022. Numbers (n) in parentheses report the corresponding total number of (observed/expected) fetal losses in the cohort.
a. The number of expected fetal losses was obtained by summing the predicted probabilities of eventual fetal loss of women in each cohort, which were estimated from the reference baseline model for each woman, based on her covariate values and gestational week of pregnancy in which she was infected (see Table S2 in Supplementary Appendix for full list of covariates and model results).
b. The top panel shows results for women infected in the respective trimester who had received no doses of COVID-19 vaccine prior to infection, and the bottom panel shows results for women who had received at least one dose of COVID-19 vaccine prior to infection. Infections were documented with PCR or (from early 2022) lateral flow tests.

**Table S9.**
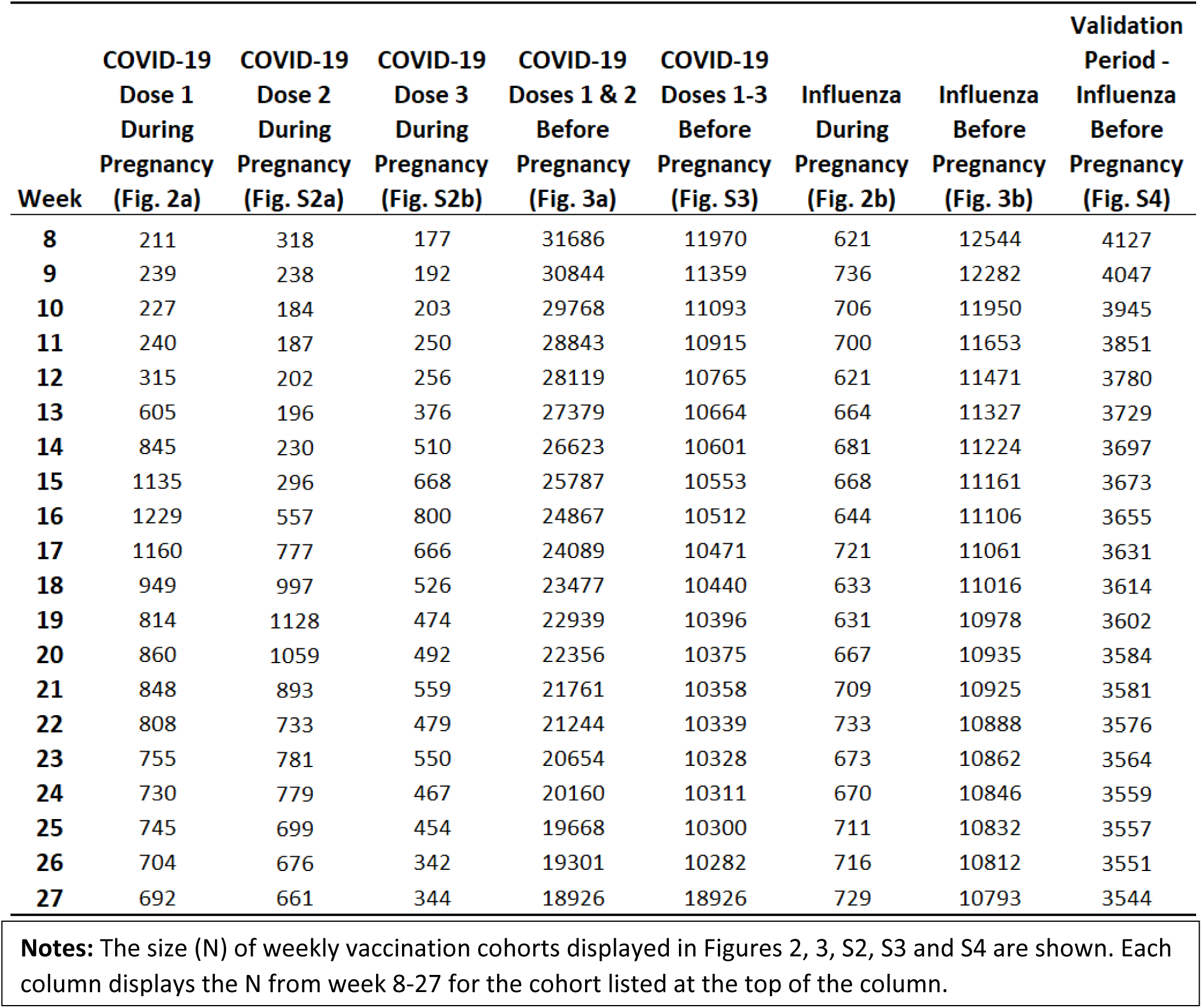
Size of Week-level Cohorts Shown in Figures.

|  | COVID-19<br>Dose 1<br>During<br>Pregnancy<br>(Fig. 2a) | COVID-19<br>Dose 2<br>During<br>Pregnancy<br>(Fig. S2a) | COVID-19<br>Dose 3<br>During<br>Pregnancy<br>(Fig. S2b) | COVID-19<br>Doses 1 & 2<br>Before<br>Pregnancy<br>(Fig. 3a) | COVID-19<br>Doses 1-3<br>Before<br>Pregnancy<br>(Fig. S3) | Influenza<br>During<br>Pregnancy<br>(Fig. 2b) | Influenza<br>Before<br>Pregnancy<br>(Fig. 3b) | Validation<br>Period -<br>Influenza<br>Before<br>Pregnancy<br>(Fig. S4) |
| --- | --- | --- | --- | --- | --- | --- | --- | --- |
| Week |  |  |  |  |  |  |  |  |
| 8 | 211 | 318 | 177 | 31686 | 11970 | 621 | 12544 | 4127 |
| 9 | 239 | 238 | 192 | 30844 | 11359 | 736 | 12282 | 4047 |
| 10 | 227 | 184 | 203 | 29768 | 11093 | 706 | 11950 | 3945 |
| 11 | 240 | 187 | 250 | 28843 | 10915 | 700 | 11653 | 3851 |
| 12 | 315 | 202 | 256 | 28119 | 10765 | 621 | 11471 | 3780 |
| 13 | 605 | 196 | 376 | 27379 | 10664 | 664 | 11327 | 3729 |
| 14 | 845 | 230 | 510 | 26623 | 10601 | 681 | 11224 | 3697 |
| 15 | 1135 | 296 | 668 | 25787 | 10553 | 668 | 11161 | 3673 |
| 16 | 1229 | 557 | 800 | 24867 | 10512 | 644 | 11106 | 3655 |
| 17 | 1160 | 777 | 666 | 24089 | 10471 | 721 | 11061 | 3631 |
| 18 | 949 | 997 | 526 | 23477 | 10440 | 633 | 11016 | 3614 |
| 19 | 814 | 1128 | 474 | 22939 | 10396 | 631 | 10978 | 3602 |
| 20 | 860 | 1059 | 492 | 22356 | 10375 | 667 | 10935 | 3584 |
| 21 | 848 | 893 | 559 | 21761 | 10358 | 709 | 10925 | 3581 |
| 22 | 808 | 733 | 479 | 21244 | 10339 | 733 | 10888 | 3576 |
| 23 | 755 | 781 | 550 | 20654 | 10328 | 673 | 10862 | 3564 |
| 24 | 730 | 779 | 467 | 20160 | 10311 | 670 | 10846 | 3559 |
| 25 | 745 | 699 | 454 | 19668 | 10300 | 711 | 10832 | 3557 |
| 26 | 704 | 676 | 342 | 19301 | 10282 | 716 | 10812 | 3551 |
| 27 | 692 | 661 | 344 | 18926 | 18926 | 729 | 10793 | 3544 |

**Notes:** The size (N) of weekly vaccination cohorts displayed in Figures 2, 3, S2, S3 and S4 are shown. Each column displays the N from week 8-27 for the cohort listed at the top of the column.

### Section S4. *Rates of Vaccination and SARS-CoV-2 Infections in the COVID-19* Period

Among the pregnancies in the *COVID-19* period included in the analysis, there are 14,111, 11,591 and 9,000 pregnancies where doses 1, 2 and 3, respectively, of the COVID-19 vaccine were received from weeks 8 to 27 (18.6%, 16.4% and 12.3%, respectively, of all pregnancies included in the *COVID-19* period). There were 39,028, 34,452 and 11,970 pregnancies where doses 1, 2 and 3, respectively, were received prior to LMP (41.4%, 36.5% and 12.7% of all pregnancies in the *COVID-19* period). Over 95% of the women vaccinated received Pfizer-BioNTech BNT162b2. Of women exposed during pregnancy, 95.6% of the first doses, 98.7% of the second doses, and 99.9% of the third doses were BNT162b2. Virtually all of the remaining women received Moderna mRNA-1273. There are 13,634 pregnancies in the *COVID-19* period where influenza vaccination was received during weeks 8-27 and 12,544 where it was administered before pregnancy but within the same influenza season (16.8% and 13.3% of all pregnancies analyzed in the *COVID-19* period, respectively).

#### SARS-CoV-2 Infections

There were 9,776 pregnancies where women had a SARS-CoV-2 infection before pregnancy and 9,073 pregnancies with an infection in weeks 8-27 (9.6% and 14.5% of pregnancies included in the *COVID-19* period, respectively), out of which 6,517 (71.8%) were infected post-vaccination. Infections were documented with PCR or (from early 2022) lateral flow tests.

**Figure S5.**
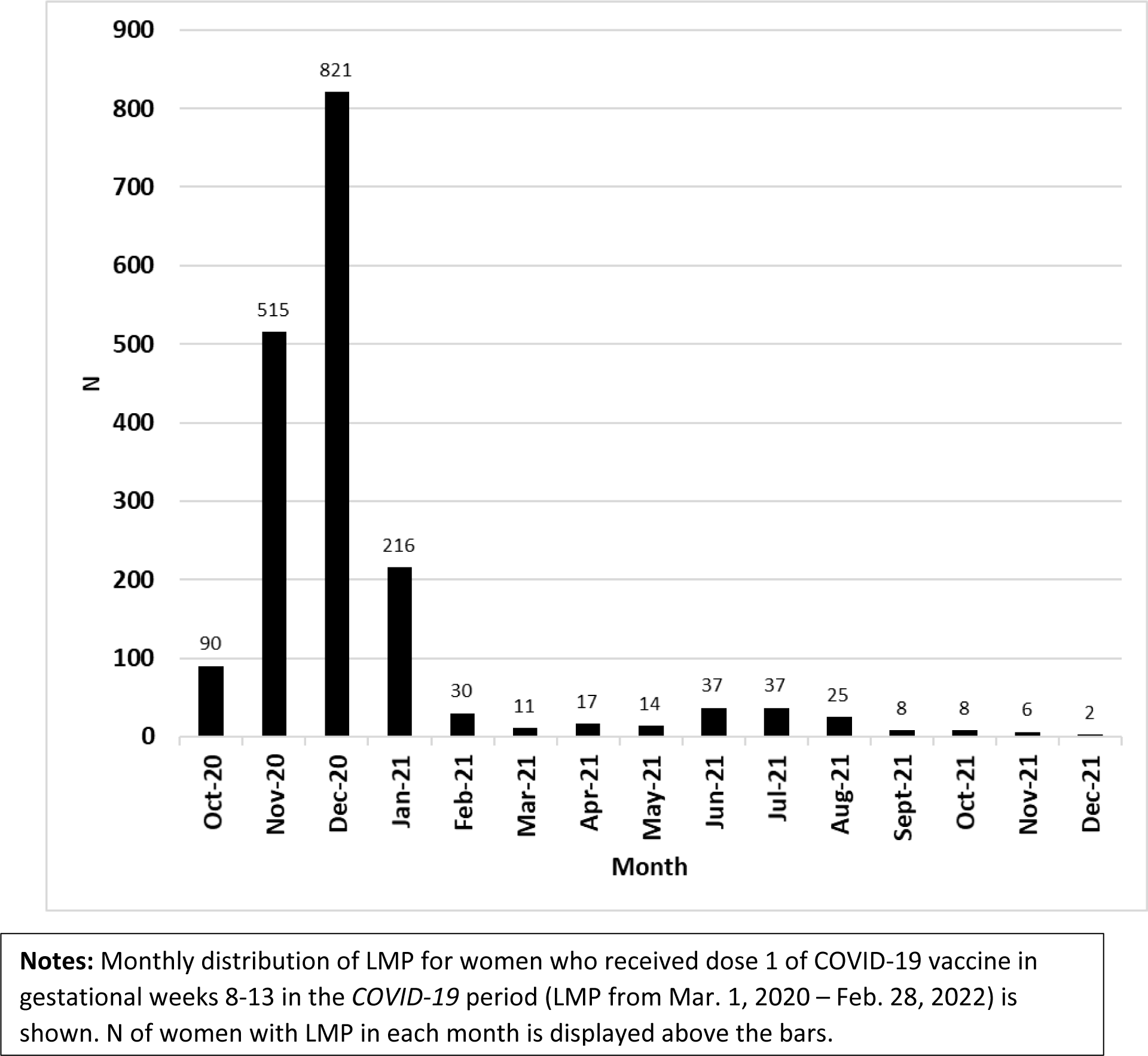
Month of LMP for Women Receiving Dose 1 of COVID-19 Vaccine in Gestational Weeks 8-13.

